# Leveraging Large Language Models for Generating Responses to Patient Messages

**DOI:** 10.1101/2023.07.14.23292669

**Authors:** Siru Liu, Allison B. McCoy, Aileen P. Wright, Babatunde Carew, Julian Z. Genkins, Sean S. Huang, Josh F. Peterson, Bryan Steitz, Adam Wright

**Author notes:** Corresponding Author: Siru Liu, PhD, Department of Biomedical Informatics, Vanderbilt University Medical Center, 2525 West End Ave #1475, Nashville, TN, 37212.

## Abstract

**Objective:** This study aimed to develop and assess the performance of fine-tuned large language models for generating responses to patient messages sent via an electronic health record patient portal.

**Methods:** Utilizing a dataset of messages and responses extracted from the patient portal at a large academic medical center, we developed a model (CLAIR-Short) based on a pre-trained large language model (LLaMA-65B). In addition, we used the OpenAI API to update physician responses from an open-source dataset into a format with informative paragraphs that offered patient education while emphasizing empathy and professionalism. By combining with this dataset, we further fine-tuned our model (CLAIR-Long). To evaluate the fine-tuned models, we used ten representative patient portal questions in primary care to generate responses. We asked primary care physicians to review generated responses from our models and ChatGPT and rated them for empathy, responsiveness, accuracy, and usefulness.

**Results:** The dataset consisted of a total of 499,794 pairs of patient messages and corresponding responses from the patient portal, with 5,000 patient messages and ChatGPT-updated responses from an online platform. Four primary care physicians participated in the survey. CLAIR-Short exhibited the ability to generate concise responses similar to provider’s responses. CLAIR-Long responses provided increased patient educational content compared to CLAIR-Short and were rated similarly to ChatGPT’s responses, receiving positive evaluations for responsiveness, empathy, and accuracy, while receiving a neutral rating for usefulness.

**Conclusion:** Leveraging large language models to generate responses to patient messages demonstrates significant potential in facilitating communication between patients and primary care providers.

## INTRODUCTION

Supported by more than $34 billion in government subsidies, the rise in adoption of electronic health records (EHRs) has led to a significant increase in the use of patient portals as a means of communication between healthcare providers and patients.[1,2] As a result, effectively managing patient messages in EHR inboxes has become an important clinical issue that needs to be addressed urgently. As an example, primary care physicians typically spend 1.5 hours per day processing approximately 150 inbox messages, continuing their work even after regular clinic hours.[3,4] This challenge is escalating due to several factors. First, the volume of patient messages is projected to grow significantly due to federal laws such as the 21st Century Cures Act which requires the instant release of test results.[5] The out-of-pocket expenses of in-person visits has also led to a preference for consultations via patient portals.[6] Finally, the pandemic prompted a 157% surge in patient messages, a trend that persisted even post-pandemic.[7] Research indicates that patients have developed an expectation for direct and prompt communication with their healthcare providers through patient portals;[6] certain time-sensitive messages, such as requests for COVID-19 antiviral medications within a five-day onset period, add to this pressure.[8] Overall, the constant influx of patient messages has evolved into a prominent stressor in clinics, particularly among primary care physicians, contributing to burnout.[9]

Large language models present a promising solution to this dilemma by enabling the automated generation of draft responses for healthcare providers. These models, trained on extensive textual data with billions of parameters, are capable of generating human-like text and performing a variety of tasks, from answering questions to summarizing and brainstorming.[10] A recent development in this domain, ChatGPT, has attracted significant attention within the medical community.[11–15] Despite not being specifically trained on medical text, ChatGPT has demonstrated impressive proficiency in medical contexts, including passing the U.S. Medical Licensing Examination (USMLE), clinical informatics board examination, and refining alert logic to improve clinical decision support (CDS).[16–18] In particular, a recent study used ChatGPT to generate responses to 195 patient questions from social media forums. The study found that ChatGPT responses outperformed those of physicians, receiving significantly higher ratings for quality and empathy.[19] This study used an “out of the box” version of ChatGPT, but it is possible to further optimize large language models’ performance on specialized domains by fine-tuning them for specific tasks.[20] For instance, a recent study utilized 100,000 patient-doctor online conversations to fine-tune the open-source Large Language Model Meta AI (LLaMA)-7B model, which showed improved performance in similarity metric (e.g., BERTScores) in comparison to ChatGPT when answering patient questions.[21] However, it is worth noting that these studies collected patient questions from online platforms, not from patient portals. A challenge with the use of the similarity metric is that it mainly measures similarity to the physician’s response, rather than accuracy or usefulness, so if the generated message is good but different from the reference response, it may score poorly.

The objectives of this study were 1) to fine-tune a large language model locally using messages and healthcare provider responses from the patient portal, and 2) to assess the generated responses from the fine-tuned model and compare them to actual provider responses and generated responses from ChatGPT3.5 and ChatGPT4. Our key advantages over prior studies are 1) our use of actual patient portal messages, 2) development of a custom model for patient message-answering and 3) scoring of responses by blinded physicians rather than similarity metrics like BERTScore.

## METHODS

### Data Collection and Preprocessing

We conducted this project at Vanderbilt University Medical Center (VUMC), a large healthcare system in the Southeastern United States using the Epic (Epic Systems Co., Verona, WI) EHR. We extracted patient messages sent to adult primary care providers along with corresponding responses from January 1, 2022 until March 7, 2023 from VUMC’s clinical data warehouse. When multiple messages were sent by a patient or a provider prior to receiving a response, we combined the messages into one. Patient messages and responses from January 1, 2022 to Feb 28, 2023 were used to develop models. To remove PHI and de-identify our dataset, we used an automated deidentification pipeline – Stanford & Penn & The Medical Imaging Data Resource Center (MIDRC) Deidentifier.[22] For instance, it replaced patient names with [PATIENT], provider names with [HCW], and telephone numbers with [PHONE].

To augment the local dataset, we randomly selected 5,000 patient questions and physician responses from an open-source dataset (including 200,000 real conversations between patients and providers on an online platform).[21] We then applied the OpenAI API (gpt-3.5-turbo) to improve the original responses into informative paragraphs with empathy and professionalism and prioritize the patient’s well-being and comfort throughout the response as a third source (Figure 1). An example of the updated response is shown in Figure 2. In our prompts, we emphasized the role by using the phase “imagine that you are a primary care doctor” to avoid GPT declining to answer medical questions. Full text of prompts is provided in Appendix 1.

**Figure 1.**
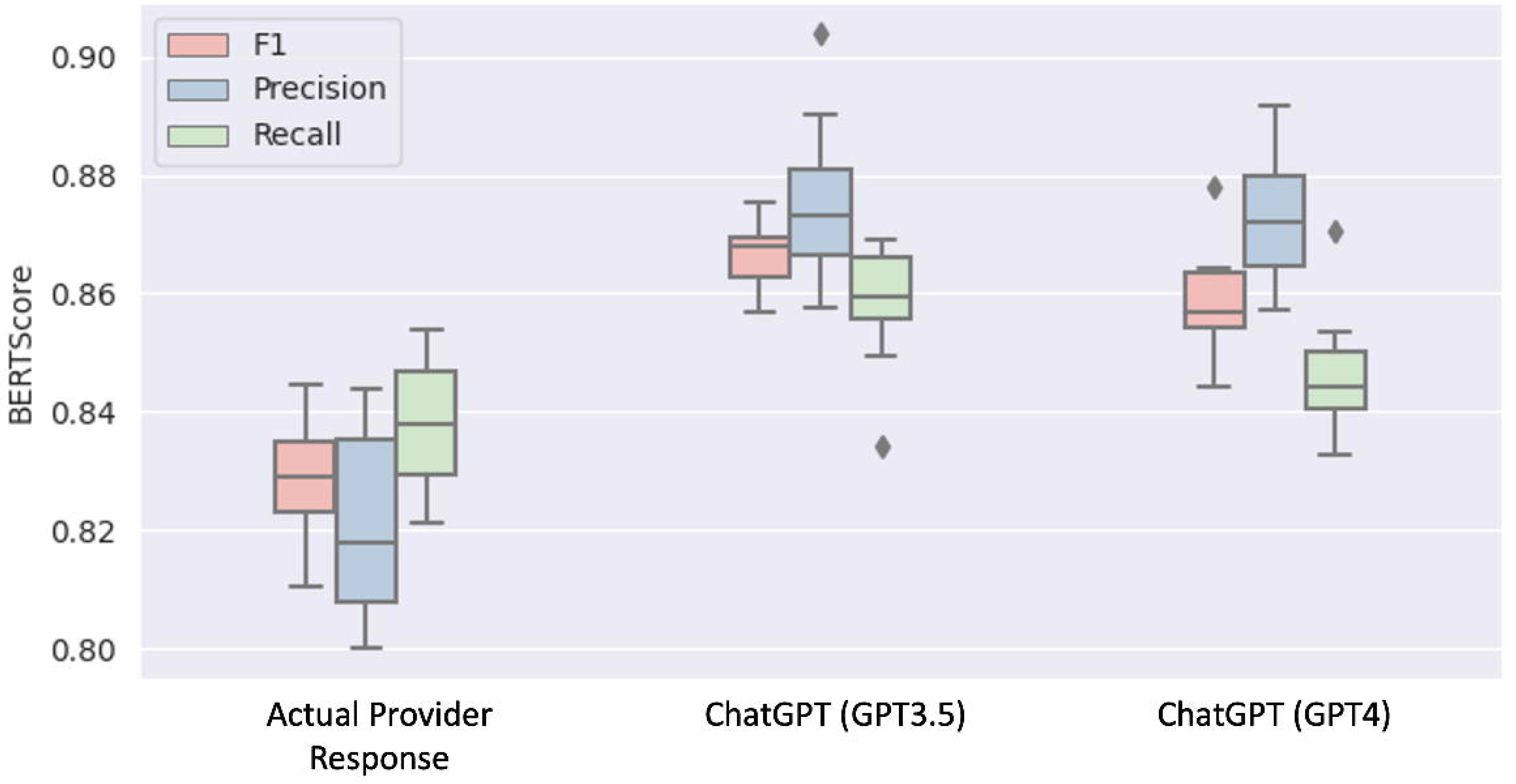
Overview of data collection, training process, and evaluation. The logos of CLAIR-Short and CLAIR-Long were generated by Midjourney.

**Figure 2.**
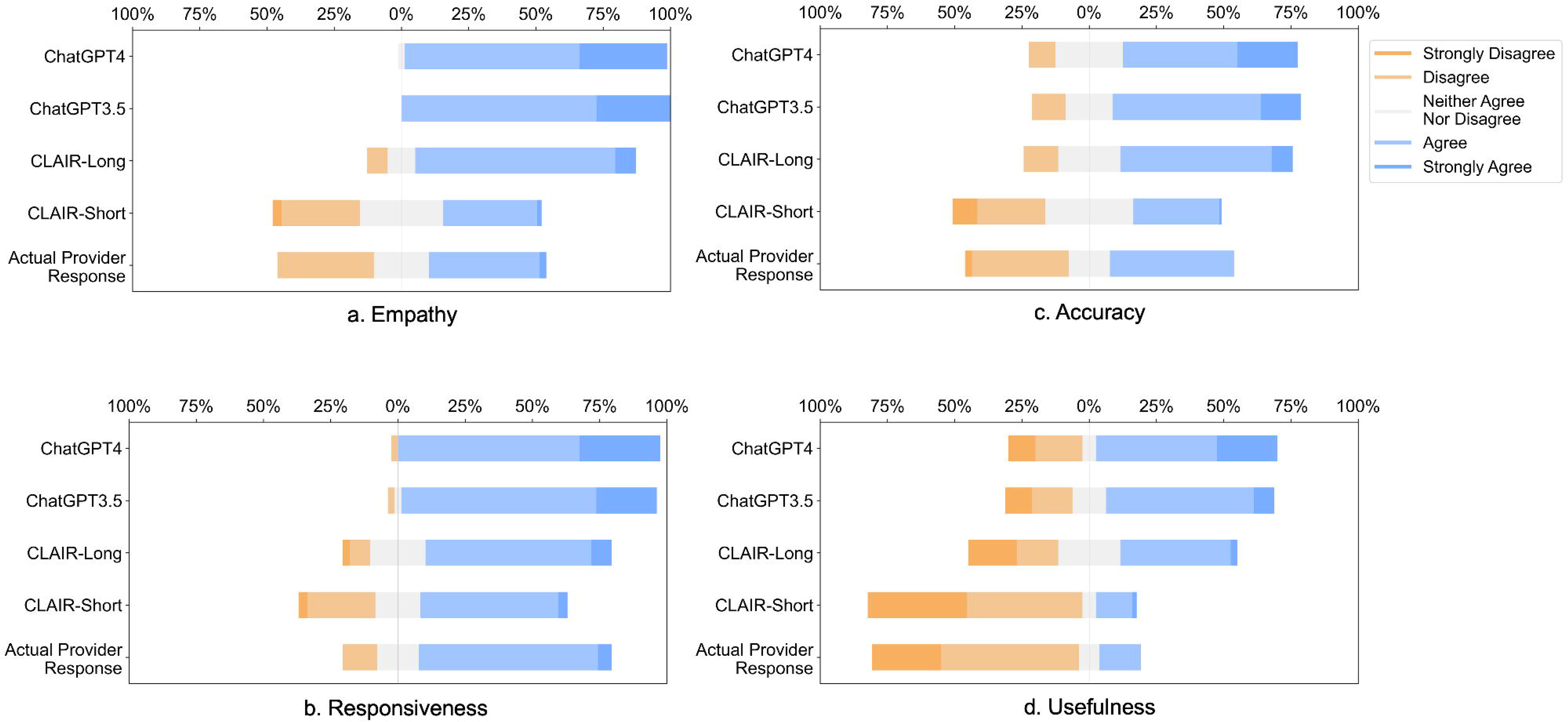
An example of updated response using OpenAI API (Turbo-3.5).

### Model Development

We developed our model using LLaMA-65B, the largest version of LLaMA models.[17,23] Leveraging low-rank adaptation, we performed supervised fine-tuning using a high-quality dataset crafted for instruction-following tasks, including data generated by GPT-4 from 52,000 prompts in Alpaca.[24,25] After gaining basic conversation capabilities, we developed two models: 1) **C**omprehensive **L**arge Language Model **A**rtificial **I**ntelligence **R**esponder (CLAIR)-Short: fine-tuned using the local dataset of patient messages and responses from VUMC, and 2) CLAIR-Long: fine-tuned using a combination of the local dataset augmented with 5,000 open-source patient questions + ChatGPT updated responses. The fine-tuning process was conducted on four A100-80G GPUs over five days with the following hyperparameters, optimizer: AdamW, batch size: 128, learning rate: 3e-4, number of epochs: 3, lora_r: 8, lora_alpha: 16, and lora_dropout: 0.05. The overview of the model development and evaluation process is shown in Figure 1.

### Evaluation Dataset

To evaluate the models, we curated a dataset from patient messages and healthcare provider responses between March 1, 2023 and March 7, 2023. We reviewed and selected 40 questions that could be answered comprehensively with minimal additional patient information and did require utilization of other tools to complete the task. A primary care physician further reviewed and ultimately selected 10 representative questions based on a patient message framework.[26] Along with removing PHI, the primary care physician created a new, rephrased message inspired by the content of the original message. The rephrased patient messages, healthcare provider responses, and corresponding categories are listed in Table 1.

**Table 1.**
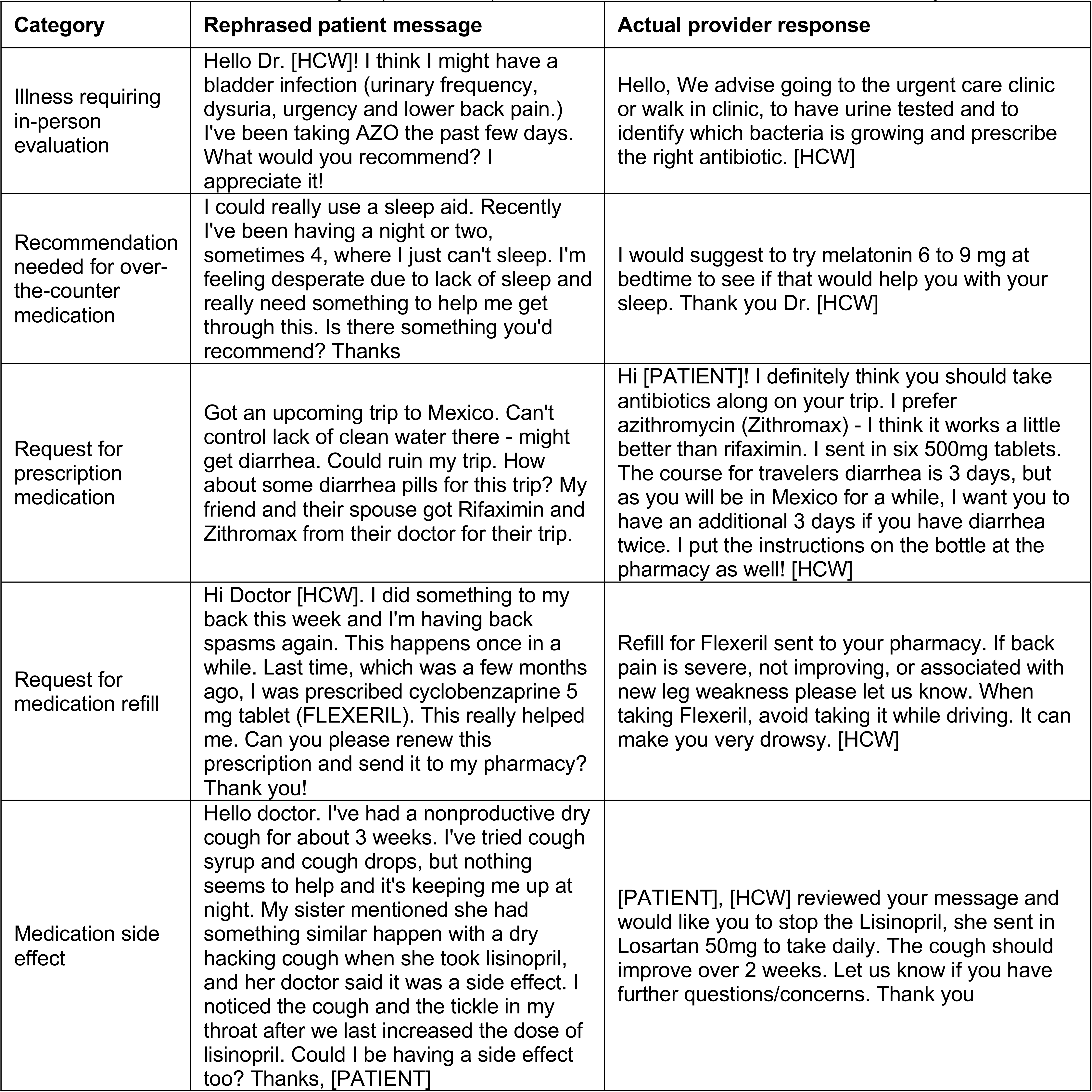
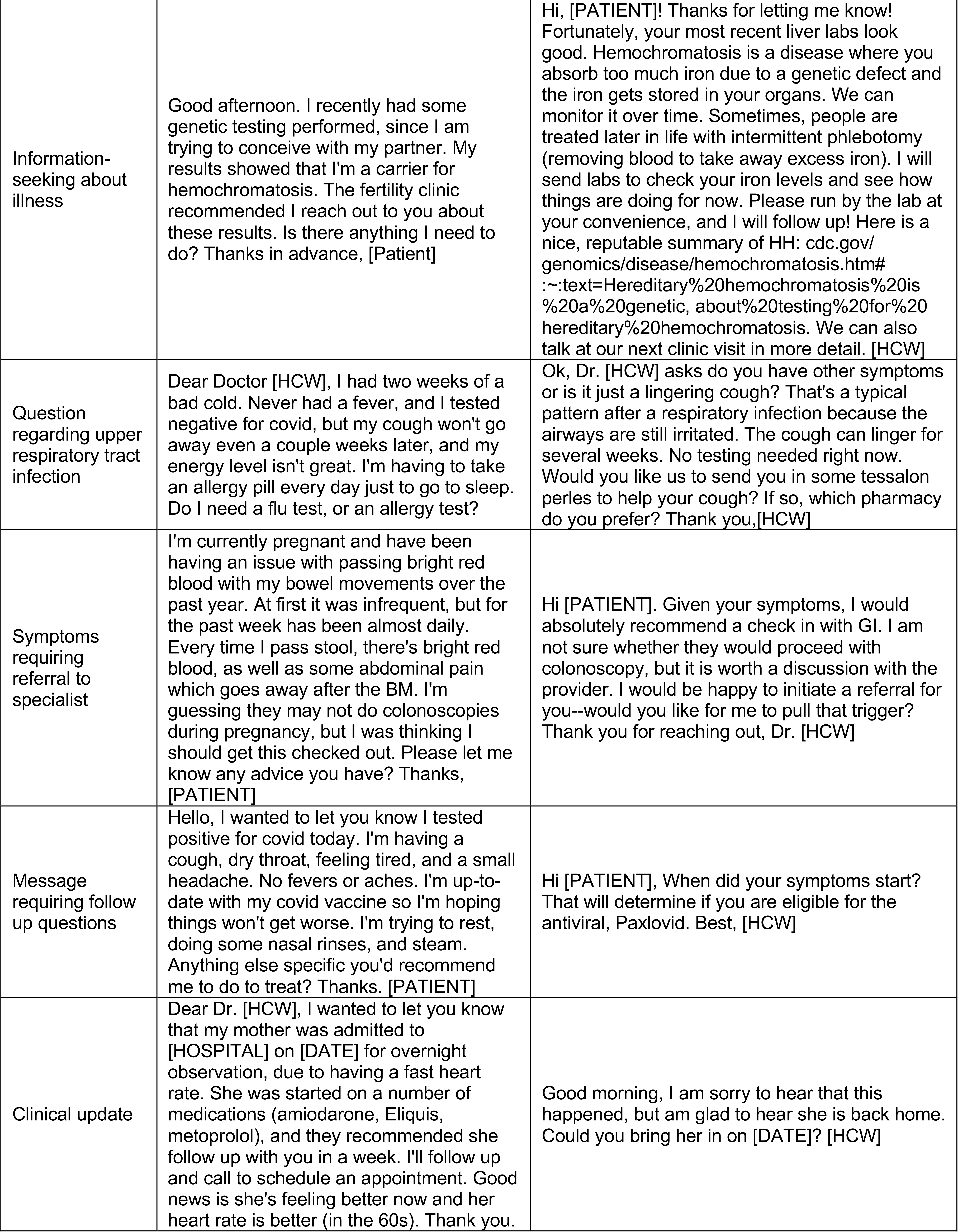
Selected patient messages (rephrased), the actual provider’s responses and categories.

We used the rephrased patient messages as input to evaluate our two fine-tuned models using a web interface developed with Gradio.[27] For comparison, we also used ChatGPT 3.5 and ChatGPT 4 to generate corresponding responses to the ten rephrased patient messages.

### Primary Care Physicians Review of Responses

For each patient message in the evaluation dataset, we randomized the order of 7 responses for review by primary care physicians: 3 from CLAIR-Short, 1 from CLAIR-Long, 1 from ChatGPT 3.5, 1 from ChatGPT 4, and 1 rephrased actual provider’s response. Participants rated each response in a survey using a 5-point Likert scale (1—strongly disagree, 5—strongly agree) from 4 perspectives: (1) **Empathy**: The answer expresses appropriate empathy given the question. (2) **Responsiveness**: The answer is responsive to the patient’s question. (3) **Accuracy**: The answer is clinically accurate, and soundly answers the patient’s question. (4) **Usefulness**: I can use it as a template to write my response to this question. Participants could also provide free-text comments for each response. Participants completed the survey using REDCap and were blinded to which model generated a given response.[28]

### Evaluation

To automatically evaluate the generated responses, we calculated BERTScore,[29] a widely used metric for evaluating generated text exhibits excellent correlation with human judgment at both sentence-level and system-level evaluations. We also computed precision, recall, and F1 scores based on BERTScore. For expert ratings, we calculated the frequencies and median and performed a Kruskal-Wallis test to compare the ratings of generated responses from different methods. To evaluate interrater reliability, we reported the intraclass correlation coefficient (ICC) and 95% confidence intervals (CIs) using a 2-way mixed-effects model.[30] The statistical analysis was performed using Python3.6.

## RESULTS

We collected 499,794 pairs of patient messages and corresponding provider responses, including interactions from 98,808 unique patients and 2,974 providers. After the removal of duplicate entries and de-identification of the data, we ended up with a final training dataset consisting of 499,286 message-response pairs. The median length was 210 characters for patient messages and 162 characters for provider responses. From the open-source dataset, median length for patient questions was 363 characters and 562 characters for provider responses. Updating the responses using the OpenAI API (Turbo-3.5), increased the length to a median 1243 characters. Figure 2 provides an example of these updated responses.

### Results of Physician Review of Responses

Four primary care physicians participated in the survey with an ICC of 0.68 [0.61, 0.74], indicating moderate reliability. We used median values of three CLAIR-Short responses as the final ratings for the CLAIR-Short model. Figure 3 displays stacked bar charts for each. Participant evaluation of ChatGPT3.5 and ChatGPT4 responses had median values leaning towards agreement in terms of empathy, responsiveness, accuracy, and usefulness, while evaluation of CLAIR-Long responses indicated agreement in empathy, responsiveness, and accuracy, but neutrality in usefulness. On the other hand, evaluation of actual provider responses and CLAIR-Short responses leaned towards disagreement in usefulness, neutrality in empathy and accuracy, and agreement in responsiveness. Pairwise comparisons of CLAIR-Long responses versus other responses revealed that CLAIR-Long responses were rated significantly higher than CLAIR-Short responses in terms of empathy (P<0.001), accuracy (P<0.001), and usefulness (P<0.001). CLAIR-Long responses were rated significantly lower than ChatGPT responses in responsiveness (P=0.005, P= 0.001). However, no statistically significant differences were observed between CLAIR-Long responses and ChatGPT3.5 or ChatGPT4 responses in terms of empathy, accuracy, and usefulness. Pairwise comparisons between other responses were notable for no statistical significance between provider’s responses and CLAIR-Short responses as well as ratings for most evaluation items between ChatGPT4 and ChatGPT3.5 responses were similar. Finally, we sought to rank the performance of each response by summing the medians of the four survey rating dimensions. Including instances of tied rankings, ChatGPT4 responses achieved the highest ranking in 6 questions, CLAIR-Long responses in 4 questions, and ChatGPT3 in 3 questions. Among 20 top-rated responses, 7 were from ChatGPT4, 5 were from ChatGPT3.5, 4 were from CLAIR-Long, 3 from CLAIR-Short, and only 1 from the actual doctor. Table 2 displays two highest-rated generated responses from each method. Medians and interquartile ranges of the survey items, detailed p values for pairwise comparisons, and median values for each response and related ranks can be found in Appendix 2.

**Figure 3.**
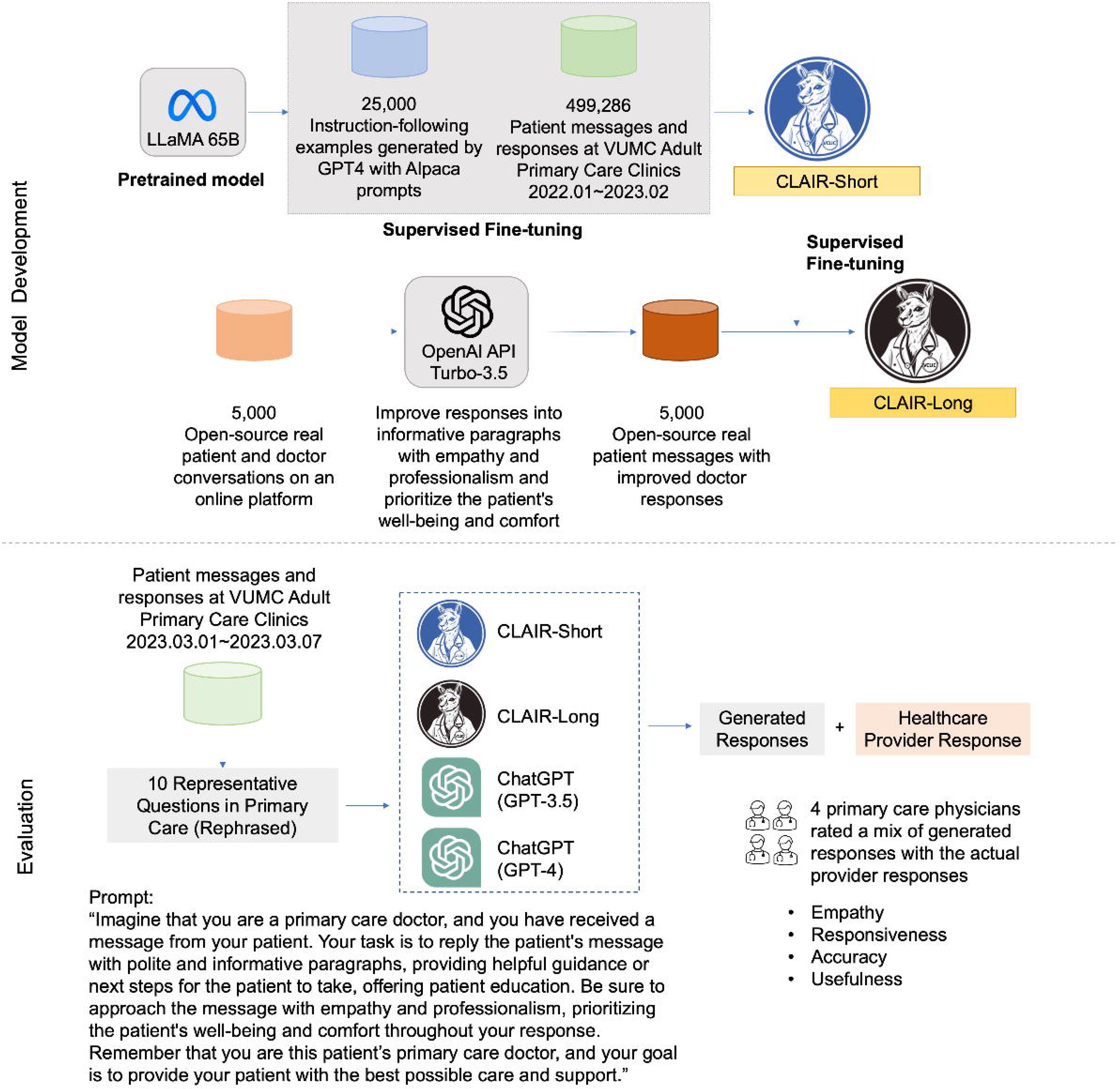
Stacked bar charts of the ratings of empathy, responsiveness, accuracy, and usefulness.

**Table 2.**
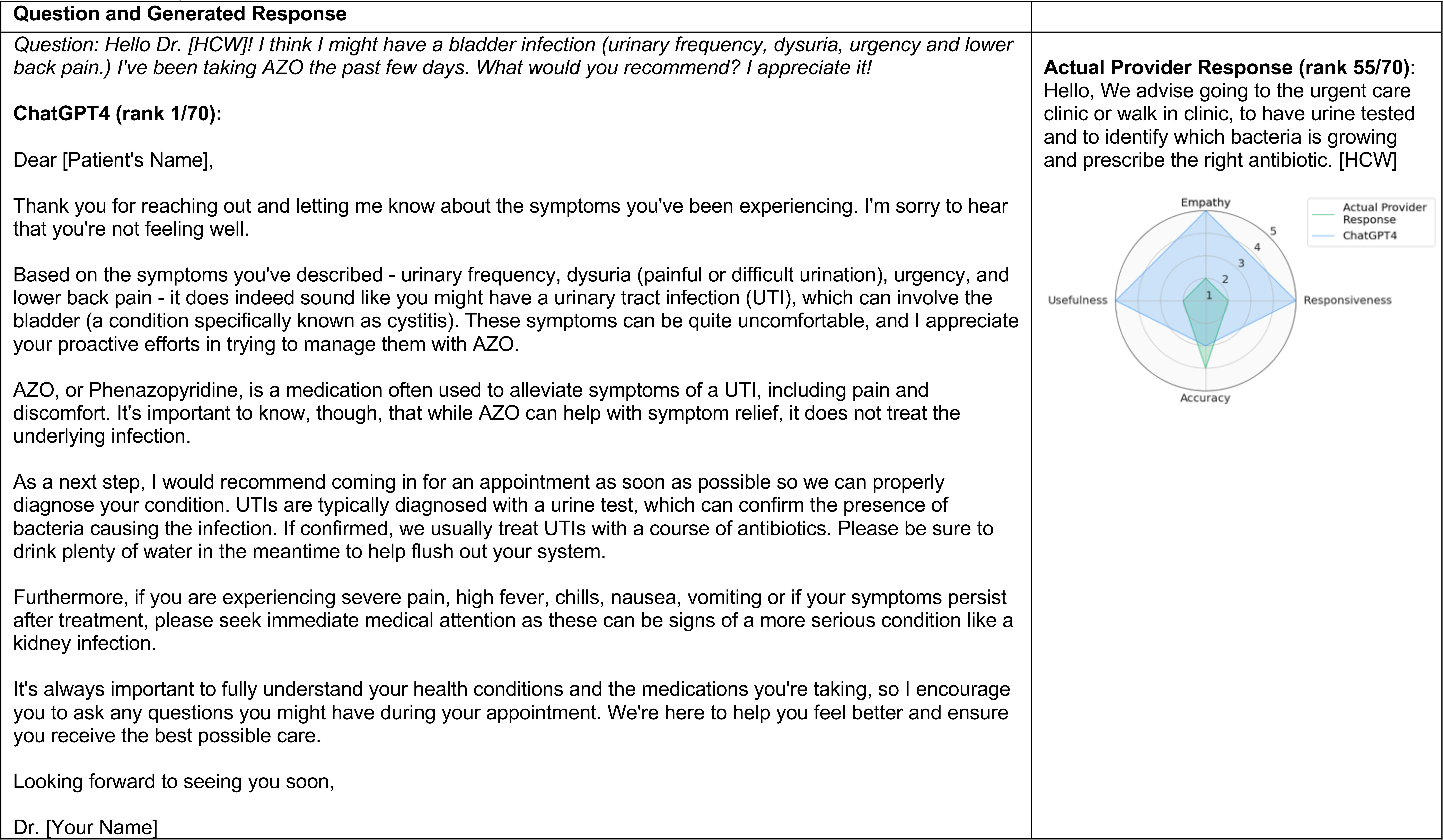
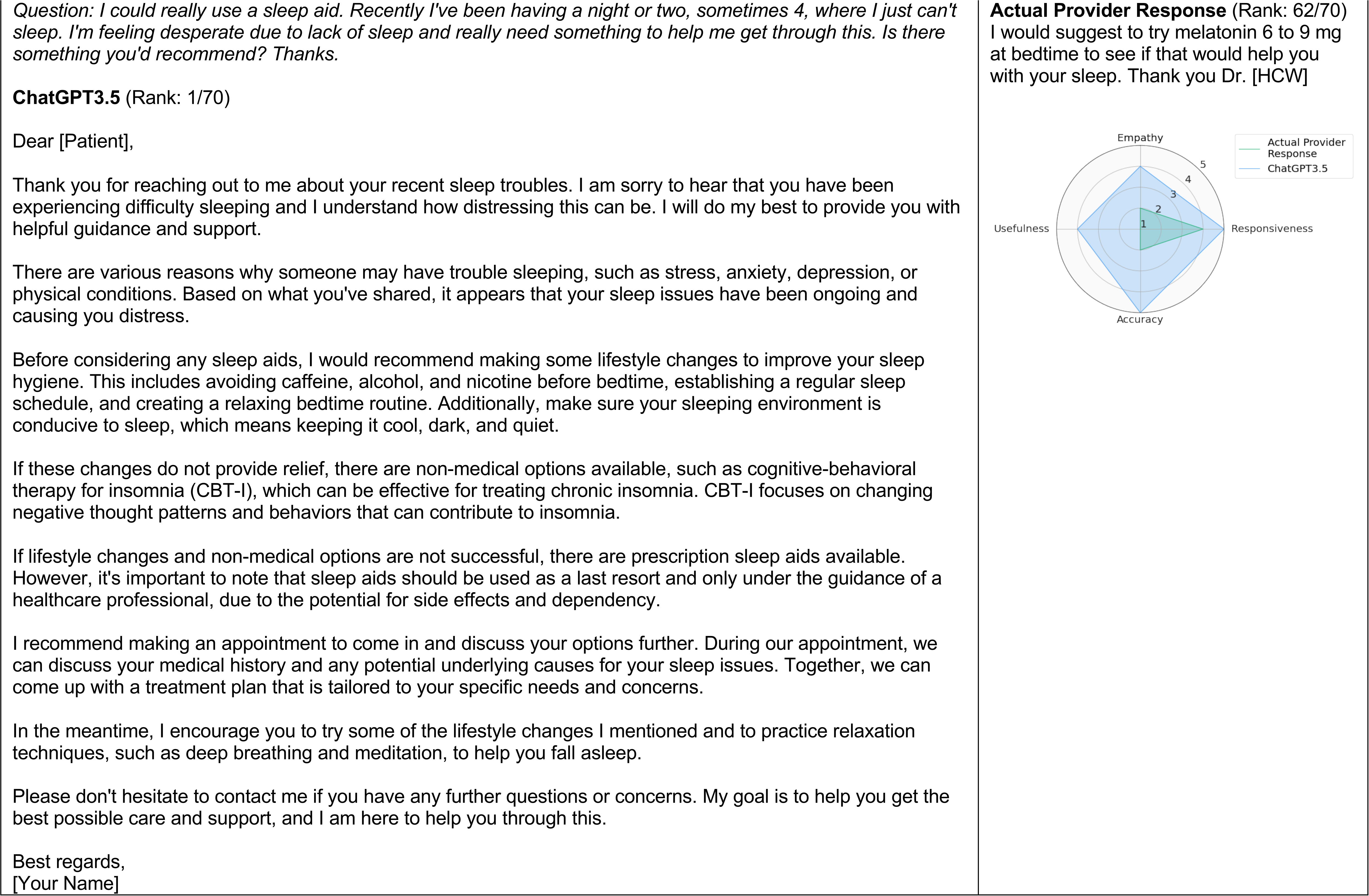
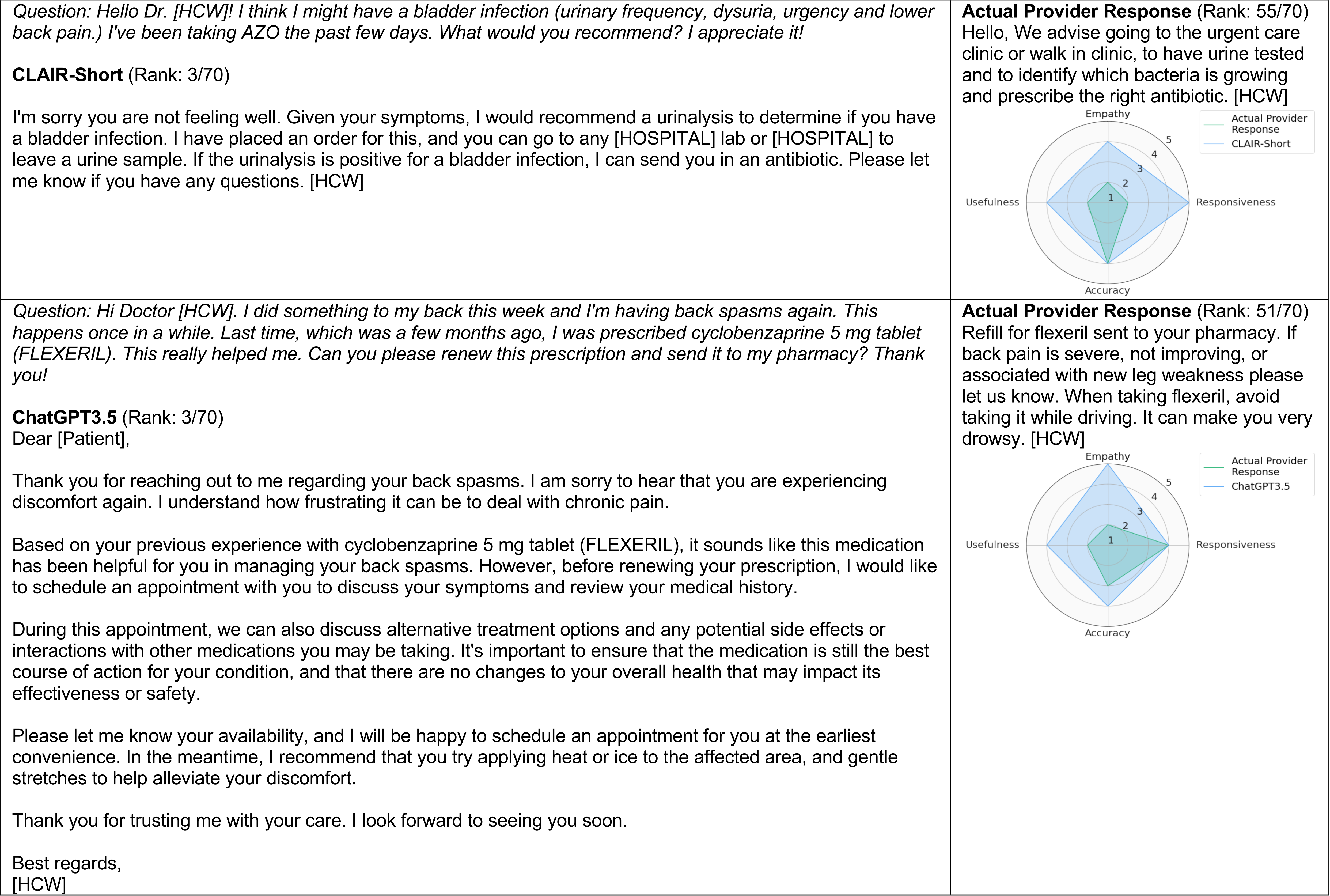
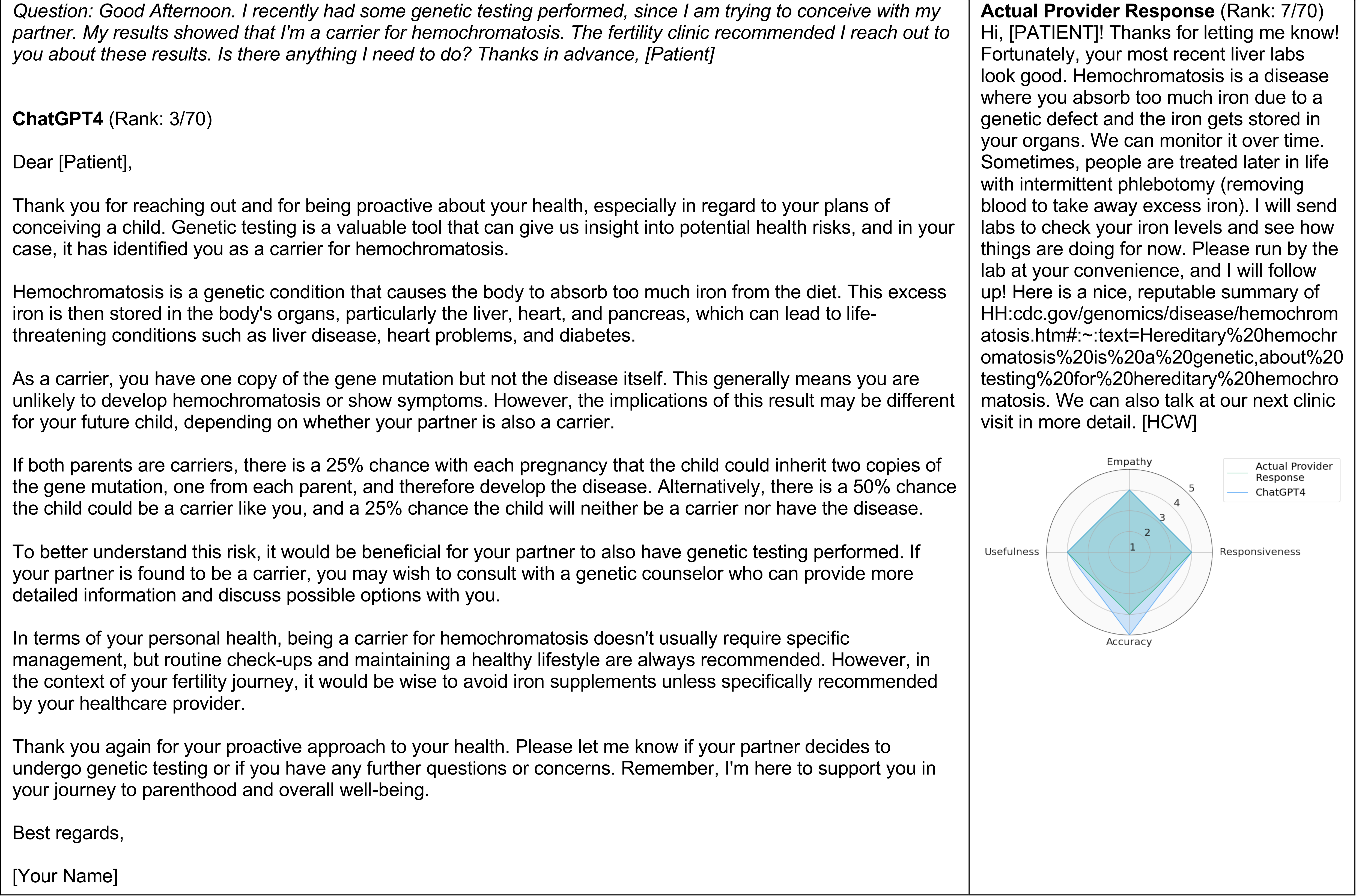
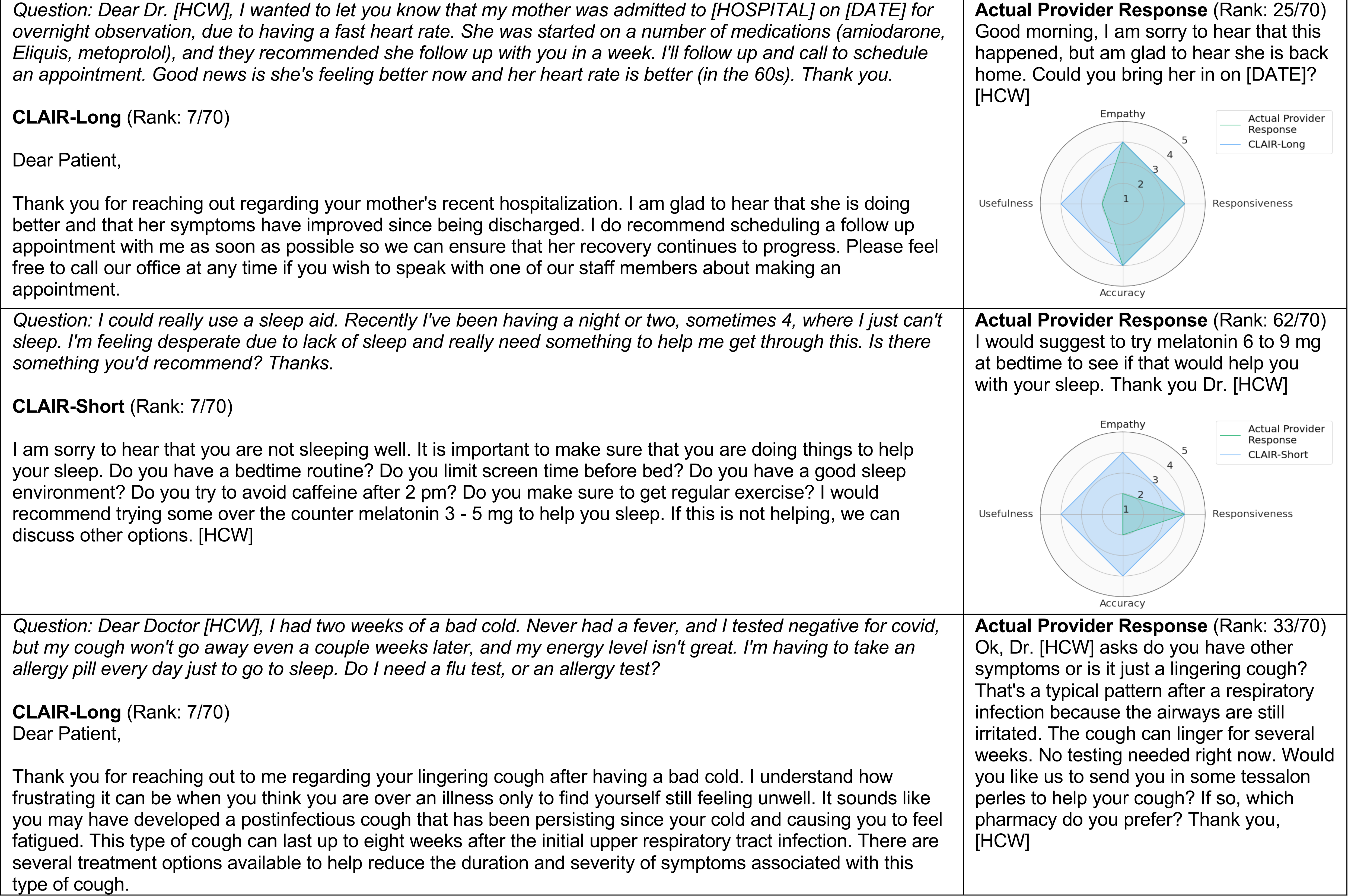
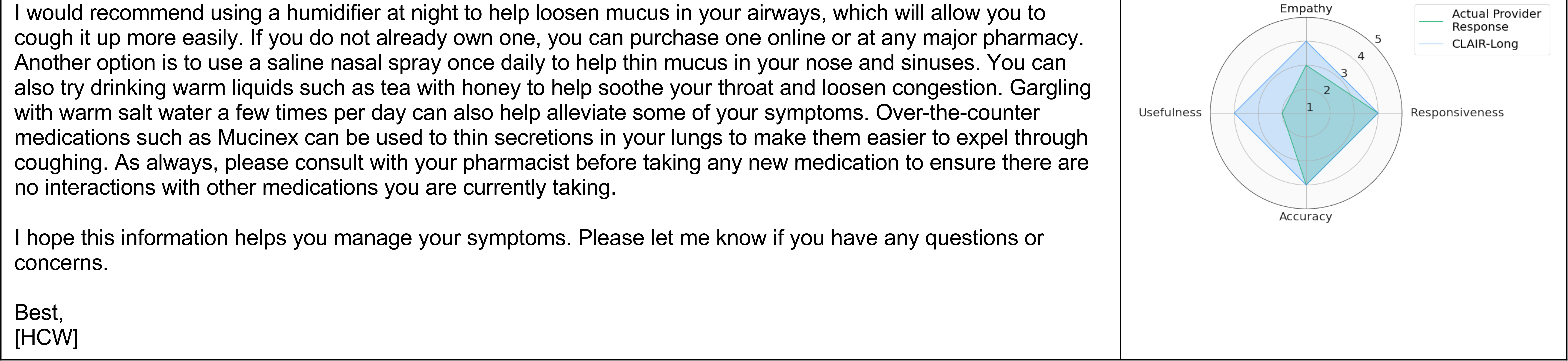
Top-rated generated responses (selected 2 responses from each generation methods). The complete set of generated responses, along with corresponding patient information and provider responses, can be found in Appendix 3.

### CLAIR-Short Generated Responses

The responses generated by CLAIR-Short had a median length of 200 characters. The ratings of these responses varied across different questions. When comparing the best generated responses from three rounds in each question with the actual provider responses, CLAIR-Short outperformed the providers in all questions except for Q6 (hemochromatosis question), where the provider responses received higher ratings. For instance, in the first scenario (Q1), where the patient had been taking AZO for several days without relief and suspected a bladder infection, the healthcare provider recommended visiting an urgent care or walk-in clinic for a urinalysis and a suitable antibiotic prescription. The CLAIR-Short generated response also suggested a urinalysis but expressed empathy. Moreover, it mentioned that a lab order had been placed, provided information on where the patient should go to provide the sample, and outlined the subsequent steps (antibiotic prescription) if the test came back positive. Reviewers also suggested that mentioning AZO could further enhance this generated response. In the second scenario (Q2), which involved a patient experiencing sleep difficulties for four consecutive days and seeking assistance, the physician suggested trying melatonin at a dose of 6 to 9 mg. The CLAIR-Short generated response displayed empathy, inquired about the patient’s sleep problems in detail, and recommended a different dose of melatonin: 3 to 5 mg. Reviewers favored the generated response and suggested that it could be further improved by discussing sleep hygiene more comprehensively. One reviewer noted a preference for discussing sleep aids with the patient before prescribing and expressed concern about the high dose mentioned in the physician’s response. Another similar scenario was presented in Q7, where a patient had a lingering cough after a cold and inquired about flu or allergy testing. The provider response requested additional information about the symptoms and offered Tessalon Perles as a cough treatment. The CLAIR-Short response included a series of follow-up questions (e.g., regarding over-the-counter cough medications, Mucinex usage, shortness of breath or chest pain, dizziness or weakness). The reviewers considered this response to be reasonable and valuable, suggesting that it could be sent to patients as an automated preliminary request for addition information before message was actually received by a provider or care team.

The average BERTScore metrics for the generated responses from CLAIR-Short, in comparison to the actual provider’s responses, were as follows: precision of 0.87±0.02, recall of 0.84±0.03, and F1 score of 0.85±0.02. The boxplot is in Figure 4.

**Figure 4.**
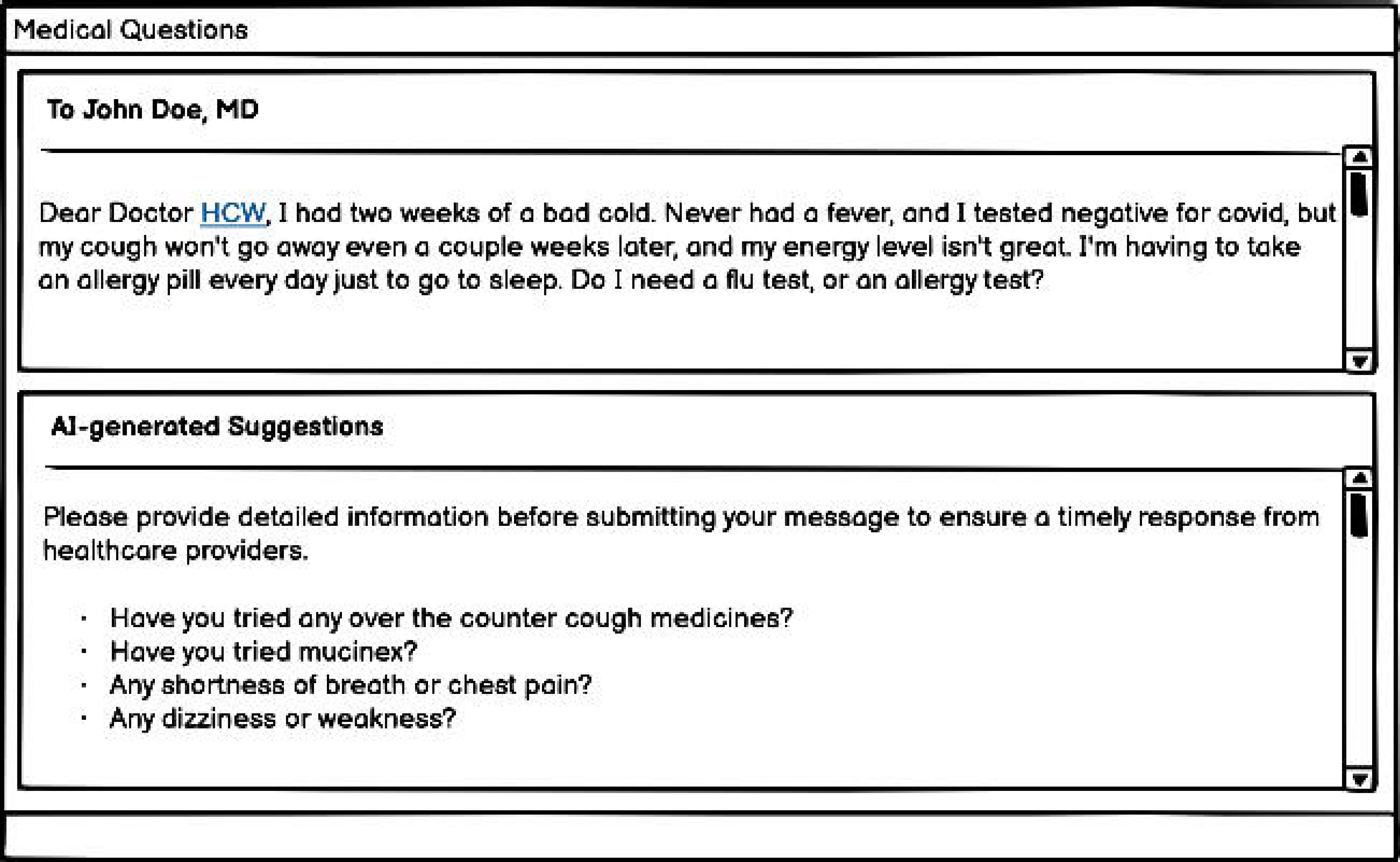
The boxplot comparing BERTScore values of generated responses from CLAIR-Short to actual provider responses.

### CLAIR-Long, ChatGPT3.5, and ChatGPT4 Generated Responses

The median length of responses generated from CLAIR-Long, ChatGPT3.5, and ChatGPT4 were 1593, 1591, and 2025 characters, respectively. In Q1, all generated responses advised patients to seek immediate medical attention and explained why their previous medication, AZO, was not sufficient for treatment. The responses from ChatGPT3.5 emphasized the importance of urine testing for diagnosis and the consideration of antibiotics based on the test results. Additionally, ChatGPT4 responses mentioned symptoms of kidney infection, urging patients to watch out for them. On the other hand, CLAIR-Long suggested evaluation at a walk-in clinic and provided a link to relevant information about urinary tract infections (UTIs). The reviewers noted that this question might require more information, such as whether the patient is pregnant. They also mentioned that UTIs involving only the bladder don’t necessarily require an appointment and can be addressed through the patient portal, while the patient’s back pain could be a symptom of a kidney infection. Another point raised was that the responses generated by ChatGPT were considered too lengthy and required a relatively high reading level. One reviewer believed that the CLAIR-Long response was the best response, while another reviewer felt it was more suitable as a nurse-directed protocol. In Q2 (sleep aid request), CLAIR-Long generated responses asked specific questions to gather more information about the patients’ symptoms, triggers, and past experiences. One reviewer noted that this response assumed that insomnia is solely caused by stress. Another reviewer mentioned that it may contain an excessive amount of empathy. On the other hand, the ChatGPT3.5 response received feedback as being highly accurate with a suggestion to make it more concise. The ChatGPT4 response received feedback suggesting that it could serve as a good template after incorporating low-risk medications and making it more concise. Generated responses are listed in Appendix 3.

Using the actual healthcare provider responses as the reference dataset, the BERTScore values for CLAIR-Long generated responses were: Precision: 0.82±0.02, Recall: 0.84±0.01, F1: 0.83±0.01. The BERTScore values of ChatGPT3.5 and ChatGPT4 generated responses compared with the CLAIR-Long generated response were Precision: 0.88±0.01, Recall: 0.86±0.01, F1: 0.87±0.01, and Precision: 0.87±0.01, Recall: 0.85±0.01, F1: 0.86±0.01, respectively. The boxplot is shown in Figure 5.

**Figure 5.**
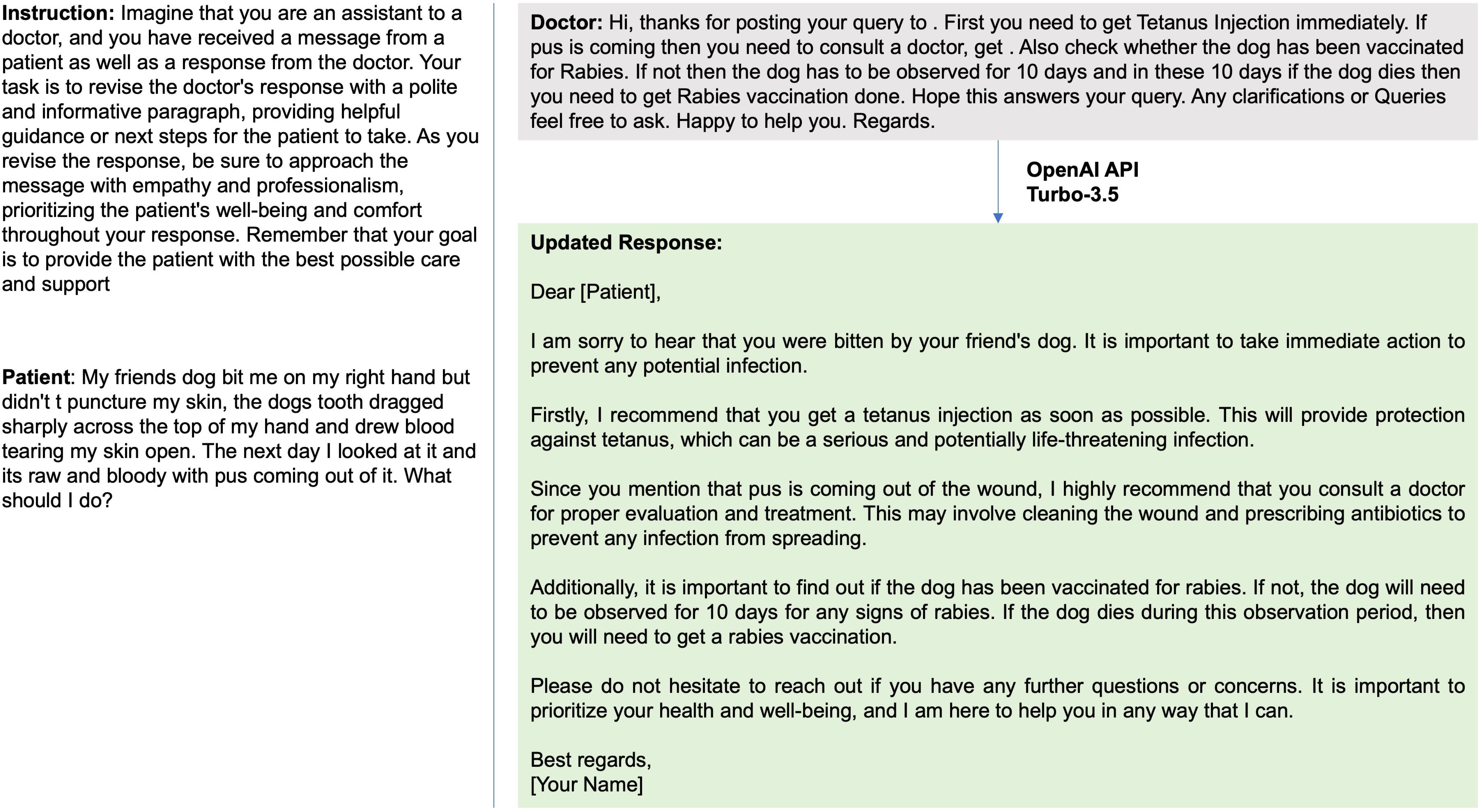
Boxplot of BERTScore of the generated responses from CLAIR-Long compared with the responses from actual providers, ChatGPT(GPT3.5), and ChatGPT(GPT4).

## DISCUSSION

In this study, we utilized GPT4 instruction data to train LLaMA-65B and developed two models for responding to patient messages. The first model, CLAIR-Short, was developed using patient messages with responses from primary care providers at VUMC. The second model, CLAIR-Long was augmented with an open-source dataset and OpenAI GPT3.5. We mixed generated responses from CLAIR-Short and CLAIR-Long with actual provider responses as well as responses from non-specialized large language models - ChatGPT3.5 and ChatGPT4. Primary care physicians evaluated these responses in terms of empathy, responsiveness, accuracy, and usefulness. The results indicated that responses generated by ChatGPT models achieved highest ratings, followed by responses generated by CLAIR-Long, both of which outperformed CLAIR-Short and the doctor’s responses significantly. In addition, we provided a set of typical patient messages and provider responses for future evaluation of response generation models in the patient portal.

### Benefits of Fine-Tuning

Although ChatGPT-generated responses received highest ratings on average, fine-tuning large language models for patient responses offers several benefits. Firstly, the fine-tuned model generates concise responses with a distinctive voice similar to local doctors. For example, CLAIR-Short-generated responses were rated as more typical of primary care physicians as compared to ChatGPT-generated responses which experts described as robot-like. Training AI generated responses to match the syntax and tone of physician authored messages may be critical to enhance both physician acceptance and patient satisfaction were such tools applied in practice. Secondly, only hospitals collaborating with Epic and Microsoft Azure have the possibility to use large language models from Open AI with PHI, such as patient messages, in a HIPAA compliant way. Fine-tuning publicly available large language models, such as LLaMA-65B, fine-tuned on local datasets could empower any researcher within any healthcare organization to do work in this area, regardless of external partnerships. Compared with CLAIR-Short’s performance limited by the quality of local data, our CLAIR-Long generated responses improved significantly by using an open-source dataset augmented with OpenAI GPT3.5. Experts generally expressed positive views on the responsiveness, empathy, and accuracy of CLAIR-Long responses, while maintaining a neutral stance on usefulness. Therefore, combining the local patient messages dataset with an augmented open-source dataset allowed effective fine-tuning of the large language model, generating responses that reflect local provider practice preferences while incorporating comprehensive information, empathy, and relevant patient education.

### ChatGPT is able to Generate Useful Draft Messages without Training on Local Data

The responses generated by ChatGPT received higher ratings compared to our fine-tuned models, which could be attributed to the superior performance of ChatGPT over the open-source large language model LLaMA. Moreover, the performance of the fine-tuned large language models depends heavily on the quality of the training dataset rather than its size.[31] In this study, the ratings for responses generated by our CLAIR-Short, which was fine-tuned solely on local data, were not significantly different from the ratings of the original physician responses across all items: empathy, responsiveness, accuracy, and usefulness. Therefore, future studies about using large language models in replying to patient messages can focus on prompt engineering, integrating large language models with EHR data and clinical knowledge dataset, helping patients draft messages, and performing patient portal tasks.

### Prompt Engineering

Prompt engineering should highlight taking the role of a primary care doctor, providing helpful guidance and patient education, and using empathy. Physician reviewers responded favorably to drafted messages that were empathetic and included patient education. Writing thorough, empathetic responses that include patient education may be beneficial for the patient but is also time-consuming, revealing a key opportunity for AI to augment clinical work.

### Clinical Context and Existing Patient-Physician Relationship

Further work is needed to incorporate patient history (e.g., medication history, diagnosis), historical conversations, and local care delivery practice preferences into prompts. During the evaluation, reviewers noted that some provider responses are based on having an established patient-provider relationship. For instance, a primary care provider may not refill Flexeril for a patient over messages alone unless they have an existing agreement and previous expectations set for short term use. In addition, using context information, we could further refine generated responses based on user types, care protocols, and patient education levels. Another finding was that some of the generated responses related to drug prescriptions did not explicitly mention specific drug names. Upon reviewing the database, we found that this communication pattern of excluding specific drug names matched with the responses from physicians, likely because the Epic EHR system had automatically generated a message to the patient earlier in the conversation which provided detailed prescription information. Therefore, when collecting training data, the prescription messages automatically generated by the system could also be collected to help improve the accuracy and completeness of the generated responses, especially when specific drug information is needed.

### Clinical Knowledge

Training, either on local datasets, or on older data may perpetuate use of out-of-date clinical guidelines. For example, in Q3 about medication request of antibiotics for traveler’s diarrhea, while the Centers for Disease Control and Prevention (CDC) Yellow Book 2024 recommends azithromycin as an alternative to fluoroquinolones, one of the generated responses still opted for ciprofloxacin. After reviewing the dataset, we found several reasons leading to this discrepancy, including provider recommending a non guideline-based antibiotic or patients explicitly requesting a specific drug based on their previous prescriptions or allergy to azithromycin. Another example is the Q9 regarding COVID-19 treatment. The doctor’s responses referred to the antiviral medication Paxlovid, which has been available from December 2021. However, responses from ChatGPT did not mention this treatment option. It might because ChatGPT only contains information from September 2021 and before. Large language models learn text patterns from the training data, which means they predict the next word based on the provided context. Therefore, if clinical guidelines change, the large language model will not update until it is retrained and, in that case, only if enough of the training text it uses reflects the new guideline. To address this, it is crucial to incorporate updated clinical guidelines into AI models by either updating the model’s knowledge, or integrating rule-based systems, or using semantic search to link with up-to-date clinical knowledge.

### Message Response Styles and Practice Patterns

Providers and care delivery systems may have different approaches, protocols, or standards of care when responding to patient messages. For example, some may attempt to diagnose and give complete treatment plans through patient portal message conversations while others prefer to have patients schedule in-person visits. Consequently, this led to different perspectives among the reviewers assessing the generated responses, and means that the definition of an ideal response is appropriately variable and organization- or provider-specific. Future tools may incorporate provider preferences into prompts, e.g., generally encouraging patients to come into clinic if treatment decisions need to be made.

### Question Generation and Chat Capability

Large language models may be useful for automatically generating questions to gather additional details from a patient before providing a manual or automated response. For some patient messages, instead of directly answering questions, our models generated a series of information-seeking questions as a reply. Further analysis of the training dataset revealed that, in clinical practice, healthcare providers often need to ask follow-up questions to gather the necessary details before communicating a finalized plan to the patient. An AI model can serve as a useful intermediary in message conversations by prompting patients with clarifying questions as they compose their messages, leveraging known strength of large language models in chat-based infrastructures. This approach could help patients provide complete information with their initial message, streamlining the subsequent conversation and minimizing back-and-forth exchange. In Figure 6, we present a prototype of an AI patient message editor as a potential integration within a patient portal interface. Future research could focus on using a similar chat-based conversation with a large language model to quickly enhance their messages by engaging with the chatbot, ensuring clarity and conciseness before sending the information to the provider.

**Figure 6.**
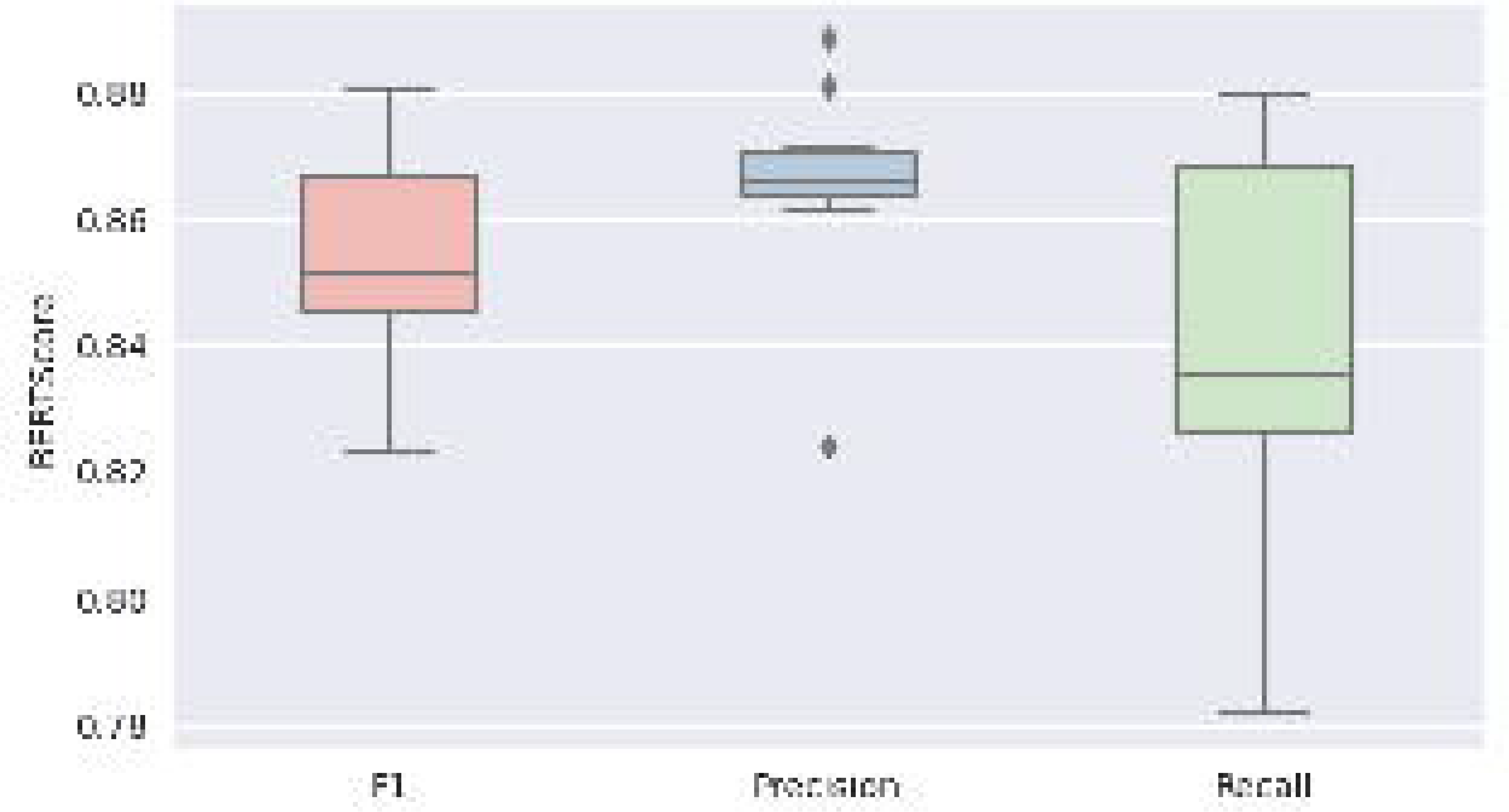
A prototype of potential implementation in of an AI Patient Message Editor in a patient portal interface.

### Patient Portal Tasks

Responses to patient portal messages often include certain tasks, like ordering tests, writing prescriptions, or scheduling appointments. Many self-service tools already exist in patient portals, such as self-scheduling or refill requests, with which patients can have their needs met in a more streamlined way without an unstructured message conversation. Furthermore, many tasks requested via messages that require care team attention can have components of the task automated, such as pending orders for medication requests or drafting letters. Future work should focus on how to use large language models to identify potential self-service redirection or automated task-completion assistance as part of the patient message response process.

### Limitations

This study has several limitations. First, the selection of patient messages to evaluate our AI models focused on single events, which might not capture the full spectrum of messages in patient portals. In reality, some patient messages require additional context, such as current medications or medical history, to provide accurate responses. Second, the models developed in this study generated responses based on previous responses stored at VUMC. Response content from a set of historical messages will not account for updates in clinical guidelines or scientific advances which occurred after the data set was created (March 7, 2023). Third, this study primarily focused on the technical feasibility of generating responses from AI models and evaluations from the physician perspective. The attitudes and preferences of patients towards these generated responses remain unknown. Future research should include qualitative studies to explore patient preferences regarding AI-generated responses and investigate any workflow issues that may arise when integrating AI-generated responses into primary care providers’ clinic work.

## CONCLUSION

Using an augmented open-source dataset can effectively improve the empathy, responsiveness, accuracy, and usefulness of responses generated by large language models fine-tuned using local data. Such open source, locally-finetuned models can perform well in generating replies to patient messages, better than actual provider responses. Generalized models like ChatGPT also outperform actual provider responses without fine-tuning on local data, as well as large language models fine-tuned with local patient message data. Locally derived models still play an important role in enabling research and clinical practice when PHI-compliant generalized large language models cannot be accessed. Further work is needed toto increase the usefulness of AI-drafted replies to patient messages, including incorporating up-to-date clinical guidelines, incorporating patient history and care-delivery context into prompts, and integrating common patient portal tasks such as pending of orders and scheduling of appointments into responses.

## CONTRIBUTORSHIP STATEMENT

SL extracted data, developed models, and drafted the work. SL, APW, ABM, and AW designed the research. SL, APW, ABM, and AW developed the questionnaire. BC, JZG, SH, and JFP participated in the questionnaire. BS provided suggestions in model development. All authors revised the draft and approved the submitted version.

## DATA AVAILABILITY STATEMENT

The updated responses based on the 5k open-source patient physician communication are available based on request.

