## Appendix for "Leveraging Large Language Models for Generating Responses to Patient Messages"

Appendix 1. Prompts.

**Prompt 1:**

Data source: Patient messages and doctor responses at VUMC Adult Primary Care Clinics

Tool: LLaMA-65B

Instruction: “Pretend you are a doctor, and you receive a message from a patient, please reply to it.”

Input: [text of patient message]

Output:[text of healthcare provider’s response].

**Prompt 2:**

Data source: Opensource patient questions and doctor responses from an online platform

Tool: OpenAI API (gpt-3.5-turbo)

The instruction was generated via ChatGPT: “Imagine that you are an assistant to a doctor, and you have received a message from a patient as well as a response from the doctor. Your task is to revise the doctor's response with a polite and informative paragraph, providing helpful guidance or next steps for the patient to take. As you revise the response, be sure to approach the message with empathy and professionalism, prioritizing the patient's well-being and comfort throughout your response. Remember that your goal is to provide the patient with the best possible care and support." The input was “Patient: [text of patient message] \n\n Doctor: [text of the healthcare provider’s response].” After receiving the updated responses, we generated a JSON file based on the following format: instruction: "Imagine that you are a primary care doctor, and you have received a message from your patient. Your task is to reply to the patient's message with polite and informative paragraphs, providing helpful guidance or next steps for the patient to take, offering patient education. Be sure to approach the message with empathy and professionalism, prioritizing the patient's well-being and comfort throughout your response. Remember that you are this patient’s doctor, and your goal is to provide your patient with the best possible care and support," input: [text of patient message], output: [text of updated response].

**Prompt 3:**

Data source: Hypothetical questions from VUMC, generated based on real questions

Tool: Chat-GPT 3.5 and 4.0

“Imagine that you are a primary care doctor, and you have received a message from your patient. Your task is to reply the patient's message with polite and informative paragraphs, providing helpful guidance or next steps for the patient to take, offering patient education. Be sure to approach the message with empathy and professionalism, prioritizing the patient's well-being and comfort throughout your response. Remember that you are this patient’s primary care doctor, and your goal is to provide your patient with the best possible care and support.”

The prompt to generate responses from CLAIR-Short and CLAIR-Long.

“Imagine that you are a primary care doctor, and you have received a message from your patient. Your task is to reply the patient's message with polite and informative paragraphs, providing helpful guidance or next steps for the patient to take, offering patient education. Be sure to approach the message with empathy and professionalism, prioritizing the patient's well-being and comfort throughout your response. Remember that you are this patient’s primary care doctor, and your goal is to provide your patient with the best possible care and support. Do not mention patients' or providers' names.”

Appendix 2. Medians and interquartile ranges for survey items using a 5-point Likert scale (1-strongly disagree, 5-strongly agree).

| | **Actual Provider Response** | **CLAIR-Short** | **CLAIR-Long** | **ChatGPT3.5** | **ChatGPT4** | **P** |
|---|---|---|---|---|---|---|
| **Empathy:** The answer expresses appropriate empathy given the question. | 3 [2, 4] | 3 [2, 4] | 4 [4, 4] | 4 [4, 5] | 4 [4, 5] | <0.001 |
| **Responsiveness**: The answer is responsive to the patient's question. | 4 [3, 4] | 4 [2, 4] | 4 [3, 4] | 4 [4, 4] | 4 [4, 5] | <0.001 |
| **Accuracy**: The answer is clinical accurate, and soundly answers the patient's question. | 3 [2, 4] | 3 [2, 4] | 4 [3, 4] | 4 [3, 4] | 4 [3, 4] | <0.001 |
| **Usefulness**: I can use it as a template to write my response to this question. | 2 [1.5, 2] | 2 [1, 2] | 3 [2, 4] | 4 [2.75, 4] | 4 [2, 4] | <0.001 |

Results of the Dunn's Test for Multiple Comparisons.

Empathy:
KruskalResult(statistic=101, P<0.001)

| | Actual Provider Response | CLAIR-Short | CLAIR-Long | ChatGPT3.5 | ChatGPT4 |
|---|---|---|---|---|---|
| **Doctor's Response** | 1 | 0.629 | 0.001 | <0.001 | <0.001 |
| **CLAIR-Short** | 0.629 | 1 | <0.001 | <0.001 | <0.001 |
| **CLAIR-Long** | 0.001 | <0.001 | 1 | 0.022 | 0.016 |
| **ChatGPT3.5** | <0.001 | <0.001 | 0.022 | 1 | 0.909 |
| **ChatGPT4** | <0.001 | <0.001 | 0.016 | 0.909 | 1 |

Responsiveness:
KruskalResult(statistic=54.92, P<0.001)

| | Actual Provider Response | CLAIR-Short | CLAIR-Long | ChatGPT3.5 | ChatGPT4 |
|---|---|---|---|---|---|
| **Doctor's Response** | 1 | 0.048 | 0.984 | 0.005 | 0.001 |
| **CLAIR-Short** | 0.048 | 1 | 0.045 | <0.001 | <0.001 |
| **CLAIR-Long** | 0.984 | 0.045 | 1 | 0.005 | 0.001 |
| **ChatGPT3.5** | 0.005 | 0.000 | 0.005 | 1 | 0.529 |
| **ChatGPT4** | 0.001 | 0.000 | 0.001 | 0.529 | 1 |

Accuracy:
KruskalResult(statistic=38.64, P<0.001)

| | Actual Provider Response | CLAIR-Short | CLAIR-Long | ChatGPT3.5 | ChatGPT4 |
|---|---|---|---|---|---|
| **Doctor's Response** | 1 | 0.401 | 0.021 | 0.003 | 0.002 |
| **CLAIR-Short** | 0.401 | 1 | <0.001 | <0.001 | <0.001 |
| **CLAIR-Long** | 0.021 | <0.001 | 1 | 0.530 | 0.456 |
| **ChatGPT3.5** | 0.003 | <0.001 | 0.530 | 1 | 0.906 |
| **ChatGPT4** | 0.002 | <0.001 | 0.456 | 0.906 | 1 |

Usefulness:
KruskalResult(statistic=64.35, P<0.001)

| | Actual Provider Response | CLAIR-Short | CLAIR-Long | ChatGPT3.5 | ChatGPT4 |
|---|---|---|---|---|---|
| **Doctor's Response** | 1 | 0.481 | 0.007 | <0.001 | <0.001 |
| **CLAIR-Short** | 0.481 | 1 | <0.001 | <0.001 | <0.001 |
| **CLAIR-Long** | 0.007 | <0.001 | 1 | 0.182 | 0.072 |
| **ChatGPT3.5** | <0.001 | <0.001 | 0.182 | 1 | 0.638 |

| | | | | | |
|---|---|---|---|---|---|
| **ChatGPT4** | <0.001 | <0.001 | 0.072 | 0.638 | 1 |

Medians for survey items using a 5-point Likert scale (1—strongly disagree, 5—strongly agree).

| Question | Method | Accuracy (median) | Empathy (median) | Responsiveness (median) | Usefulness (median) | Rank |
|---|---|---|---|---|---|---|
| q1 | ChatGPT3.5 | 3 | 5 | 5 | 4 | 2 |
| **q1** | **ChatGPT4** | 3 | 5 | 5 | 5 | 1 |
| q1 | Actual Provider Response | 4 | 2 | 2 | 2 | 5 |
| q1 | CLAIR-Long | 4 | 4 | 4 | 3 | 3 |
| q1 | CLAIR-Short | 4 | 4 | 4 | 2 | 4 |
| **q2** | **ChatGPT3.5** | 5 | 4 | 5 | 4 | 1 |
| q2 | ChatGPT4 | 4 | 4 | 4 | 4 | 2 |
| q2 | Actual Provider Response | 2 | 2 | 4 | 1 | 5 |
| q2 | CLAIR-Long | 4 | 4 | 3 | 3 | 3 |
| q2 | CLAIR-Short | 3 | 3 | 4 | 2 | 4 |
| q3 | ChatGPT3.5 | 2 | 4 | 4 | 3 | 3 |
| q3 | ChatGPT4 | 4 | 4 | 4 | 2 | 2 |
| q3 | Actual Provider Response | 2 | 4 | 4 | 2 | 4 |
| **q3** | **CLAIR-Long** | 4 | 4 | 4 | 4 | 1 |
| q3 | CLAIR-Short | 2 | 3 | 3 | 1 | 5 |
| **q4** | **ChatGPT3.5** | 4 | 5 | 4 | 4 | 1 |
| q4 | ChatGPT4 | 4 | 4 | 4 | 4 | 2 |
| q4 | Actual Provider Response | 3 | 2 | 4 | 2 | 4 |
| q4 | CLAIR-Long | 4 | 4 | 4 | 3 | 3 |
| q4 | CLAIR-Short | 2 | 3 | 3 | 1 | 5 |
| q5 | ChatGPT3.5 | 4 | 4 | 4 | 3 | 2 |
| **q5** | **ChatGPT4** | 4 | 4 | 4 | 4 | 1 |
| q5 | Actual Provider Response | 4 | 2 | 3 | 1 | 4 |
| q5 | CLAIR-Long | 4 | 4 | 4 | 2 | 3 |
| q5 | CLAIR-Short | 2 | 3 | 4 | 1 | 4 |
| q6 | ChatGPT3.5 | 4 | 4 | 4 | 4 | 2 |
| **q6** | **ChatGPT4** | 5 | 4 | 4 | 4 | 1 |
| q6 | Actual Provider Response | 4 | 4 | 4 | 4 | 2 |
| q6 | CLAIR-Long | 2.5 | 3 | 4 | 1.5 | 4 |
| q6 | CLAIR-Short | 3 | 2 | 4 | 2 | 4 |
| q7 | ChatGPT3.5 | 4 | 4 | 4 | 2 | 2 |

| | | | | | | |
|---|---|---|---|---|---|---|
| q7 | ChatGPT4 | 3 | 4 | 4 | 2 | 3 |
| q7 | Actual Provider Response | 4 | 3 | 4 | 2 | 3 |
| **q7** | **CLAIR-Long** | 4 | 4 | 4 | 4 | 1 |
| q7 | CLAIR-Short | 4 | 3 | 4 | 2 | 3 |
| q8 | ChatGPT3.5 | 3 | 4 | 4 | 2 | 2 |
| **q8** | **ChatGPT4** | 3 | 4 | 4 | 3 | 1 |
| q8 | Actual Provider Response | 2 | 4 | 4 | 2 | 4 |
| q8 | CLAIR-Long | 3 | 4 | 4 | 2 | 2 |
| q8 | CLAIR-Short | 3 | 3 | 4 | 1 | 5 |
| **q9** | **ChatGPT3.5** | 4 | 4 | 4 | 4 | 1 |
| **q9** | **ChatGPT4** | 4 | 4 | 4 | 4 | 1 |
| q9 | Actual Provider Response | 4 | 3 | 4 | 2 | 4 |
| **q9** | **CLAIR-Long** | 4 | 4 | 4 | 4 | 1 |
| q9 | CLAIR-Short | 3 | 4 | 4 | 2 | 4 |
| q10 | ChatGPT3.5 | 4 | 4 | 4 | 3 | 3 |
| **q10** | **ChatGPT4** | 4 | 4 | 4 | 4 | 1 |
| q10 | Actual Provider Response | 4 | 4 | 4 | 2 | 4 |
| **q10** | **CLAIR-Long** | 4 | 4 | 4 | 4 | 1 |
| q10 | CLAIR-Short | 4 | 4 | 4 | 2 | 4 |

Appendix 3. Responses and comments.

| Method | Question | Response |
|---|---|---|
| ChatGPT4 | Question 1: Hello Dr. [HCW]! I think I might have a bladder infection (urinary frequency, dysuria, urgency and lower back pain.) I've been taking AZO the past few days. What would you recommend? I appreciate it! | Dear [Patient's Name],<br><br>Thank you for reaching out and letting me know about the symptoms you've been experiencing. I'm sorry to hear that you're not feeling well.<br><br>Based on the symptoms you've described - urinary frequency, dysuria (painful or difficult urination), urgency, and lower back pain - it does indeed sound like you might have a urinary tract infection (UTI), which can involve the bladder (a condition specifically known as cystitis). These symptoms can be quite uncomfortable, and I appreciate your proactive efforts in trying to manage them with AZO.<br><br>AZO, or Phenazopyridine, is a medication often used to alleviate symptoms of a UTI, including pain and discomfort. It's important to know, though, that while AZO can help with symptom relief, it does not treat the underlying infection.<br><br>As a next step, I would recommend coming in for an appointment as soon as possible so we can properly diagnose your condition. UTIs are typically diagnosed with a urine test, which can confirm the presence of bacteria causing the infection. If confirmed, we usually treat UTIs with a course of antibiotics. Please be sure to drink plenty of water in the meantime to help flush out your system.<br><br>Furthermore, if you are experiencing severe pain, high fever, chills, nausea, vomiting or if your symptoms persist after treatment, please seek immediate medical attention as these can be signs of a more serious condition like a kidney infection.<br><br>It's always important to fully understand your health conditions and the medications you're taking, so I encourage you to ask any questions you might have during your appointment. We're here to help you feel better and ensure you receive the best possible care.<br><br>Looking forward to seeing you soon,<br><br>Dr. [Your Name] |
| ChatGPT3 | Question 1: Hello Dr. [HCW]! I think I might have a bladder infection (urinary frequency, dysuria, urgency and lower back pain.) I've been taking AZO the past few days. What would you recommend? I appreciate it! | Dear [Patient's Name],<br><br>I'm sorry to hear that you are experiencing symptoms that suggest a bladder infection. As your primary care doctor, I am here to help you manage your health concerns and provide you with the best possible care.<br><br>Based on the symptoms you described, it is likely that you are indeed experiencing a bladder infection. The medication you have been taking, AZO, is an over-the-counter drug that can help alleviate some of the discomfort associated with bladder infections. However, it does not cure the infection itself. |

| | | |
|---|---|---|
| | | Therefore, I strongly recommend that you come in for an appointment at your earliest convenience. During the appointment, I will perform a physical examination and may order a urine test to confirm the diagnosis of a bladder infection. Depending on the severity of the infection, I may prescribe antibiotics to help clear the infection and prevent any complications.<br><br>In the meantime, it's important to continue taking AZO as directed to manage your symptoms. It's also crucial to stay hydrated by drinking plenty of water, as this can help flush out any bacteria in your urinary tract.<br><br>Bladder infections can be uncomfortable and disruptive to your daily life, so I understand how important it is to find relief as soon as possible. Please let me know if you have any further questions or concerns, and we can work together to get you feeling better.<br><br>Best regards,<br><br>[Your Name] |
| CLAIR-Long | Question 1: Hello Dr. [HCW]! I think I might have a bladder infection (urinary frequency, dysuria, urgency and lower back pain.) I've been taking AZO the past few days. What would you recommend? I appreciate it! | Dear Patient,<br><br>Thank you for reaching out to me regarding your symptoms of potential urinary tract infection. Based on your symptoms, I do believe that you may be experiencing a UTI and would suggest that you visit one of our walk-in clinics to receive treatment today. While over-the-counter medications such as Azo can help ease some of your discomfort, they will not treat the bacterial infection itself. It is important to seek evaluation by a medical provider to ensure that you receive the appropriate antibiotic treatment for your infection. You can find the location of your nearest Walk-In Clinic, at https://www.vanderbilthealth.org/walk-in-clinics/.<br><br>I have also attached a link to our website below which provides additional information about UTIs and their treatment. Please let me know if you have any other questions or concerns, or if there is anything else that I can help you with. I wish you a speedy recovery and feel better soon. |
| CLAIR-Short | Question 1: Hello Dr. [HCW]! I think I might have a bladder infection (urinary frequency, dysuria, urgency and lower back pain.) I've been taking AZO the past few days. What would you recommend? I appreciate it! | I'm sorry you are not feeling well. Given your symptoms, I would recommend a urinalysis to determine if you have a bladder infection. I have placed an order for this, and you can go to any [HOSPITAL] lab or [HOSPITAL] to leave a urine sample. If the urinalysis is positive for a bladder infection, I can send you in an antibiotic. Please let me know if you have any questions. [HCW] |
| Actual Provider Response | Question 1: Hello Dr. [HCW]! I think I might have a bladder infection (urinary frequency, dysuria, urgency and lower back pain.) I've been taking AZO the past few days. What would you recommend? I appreciate it! | Hello, We advise going to the urgent care clinic or walk in clinic, to have urine tested and to identify which bacteria is growing and prescribe the right antibiotic. [HCW] |
| CLAIR-Short | Question 1: Hello Dr. [HCW]! I think I might have a bladder infection (urinary frequency, dysuria, urgency and lower back pain.) I've been taking AZO the past few days. What would you recommend? I appreciate it! | I am so sorry you are not feeling well. I would recommend going to a walk in clinic to be evaluated. A provider there will be able to order a urinalysis and treat you appropriately. Please let me know if you have any further questions. Thanks, [HCW] |

| | | |
|---|---|---|
| CLAIR-Short | Question 1: Hello Dr. [HCW]! I think I might have a bladder infection (urinary frequency, dysuria, urgency and lower back pain.) I've been taking AZO the past few days. What would you recommend? I appreciate it! | I would need to see you to assess and order a urinalysis to check for infection. Please call [PHONE] to schedule an appointment. Thanks. |
| ChatGPT4 | Question 10: Dear Dr. [HCW], I wanted to let you know that my mother was admitted to [HOSPITAL] on [DATE] for overnight observation, due to having a fast heart rate. She was started on a number of medications (amiodarone, Eliquis, metoprolol), and they recommeded she follow up with you in a week. I'll follow up and call to schedule an appointment. Good news is she's feeling better now and her heart rate is better (in the 60s). Thank you. | Dear [Patient's Name],<br><br>Thank you for letting me know about your mother's recent hospitalization. I'm relieved to hear she's feeling better and that her heart rate has stabilized. It's certainly important for us to follow up closely on this matter.<br><br>Amiodarone, Eliquis, and Metoprolol are medications often used to manage heart rhythm abnormalities and to reduce the risk of blood clots or stroke. However, like all medications, they can have side effects, and their usage requires careful monitoring. When your mother comes in for her appointment, we'll discuss these medications in detail, ensuring she understands their purpose, potential side effects, and any necessary lifestyle modifications.<br><br>I'm glad to hear that you will be scheduling an appointment for her. It's important that we monitor her condition closely, especially in these initial stages of her new treatment regimen. Please bring along any discharge instructions or documents from the hospital, as these will be helpful in our discussion.<br><br>While we wait for the appointment, please keep a watchful eye on your mother's symptoms. If she experiences any side effects from the medication, such as dizziness, unusual fatigue, shortness of breath, or if her heart rate becomes unusually high or low again, please seek immediate medical attention.<br><br>It's great to hear that you're actively involved in your mother's care. This can be an overwhelming time, and your support is invaluable. If you or your mother have any questions or concerns before the appointment, please don't hesitate to contact our office. We're here to provide the best care and support possible for your mother.<br><br>Kind regards,<br>Dr. [HCW] |
| CLAIR-Long | Question 10: Dear Dr. [HCW], I wanted to let you know that my mother was admitted to [HOSPITAL] on [DATE] for overnight observation, due to having a fast heart rate. She was started on a number of medications (amiodarone, Eliquis, metoprolol), and they recommeded she follow up with you in a week. I'll follow up and call to schedule an appointment. Good news is she's feeling better now and her heart rate is better (in the 60s). Thank you. | Dear Patient,<br><br>Thank you for reaching out regarding your mother's recent hospitalization. I am glad to hear that she is doing better and that her symptoms have improved since being discharged. I do recommend scheduling a follow up appointment with me as soon as possible so we can ensure that her recovery continues to progress. Please feel free to call our office at any time if you wish to speak with one of our staff members about making an appointment. |

| | | |
|---|---|---|
| ChatGPT3 | Question 10: Dear Dr. [HCW], I wanted to let you know that my mother was admitted to [HOSPITAL] on [DATE] for overnight observation, due to having a fast heart rate. She was started on a number of medications (amiodarone, Eliquis, metoprolol), and they recommeded she follow up with you in a week. I'll follow up and call to schedule an appointment. Good news is she's feeling better now and her heart rate is better (in the 60s). Thank you. | Dear [Patient],<br><br>Thank you for letting me know about your mother's recent hospitalization. I am sorry to hear that she had to go through that, but I am glad to hear that she is feeling better now and that her heart rate has improved.<br><br>Based on the medications you mentioned, it sounds like your mother may have been diagnosed with an arrhythmia, which is an abnormal heart rhythm. Amiodarone is a medication used to treat certain types of arrhythmias, while Eliquis is an anticoagulant medication used to prevent blood clots in patients with atrial fibrillation, which is a common type of arrhythmia. Metoprolol is a medication used to slow down the heart rate and reduce blood pressure.<br><br>It is important that your mother continues to take her medications as prescribed and follows up with me as recommended by the hospital. During her follow-up appointment, we will review her medications and discuss any potential side effects or interactions. We will also evaluate her response to treatment and make any necessary adjustments to her medications.<br><br>In the meantime, I recommend that your mother monitors her symptoms and alerts us immediately if she experiences any concerning symptoms such as chest pain, shortness of breath, or fainting. It is also important that she stays hydrated, eats a healthy diet, and gets enough rest.<br><br>Please feel free to call my office to schedule a follow-up appointment at your earliest convenience. If you have any questions or concerns, please do not hesitate to reach out to me. Thank you for entrusting me with your mother's care.<br><br>Best regards,<br>[Your Name] |
| Actual Provider Response | Question 10: Dear Dr. [HCW], I wanted to let you know that my mother was admitted to [HOSPITAL] on [DATE] for overnight observation, due to having a fast heart rate. She was started on a number of medications (amiodarone, Eliquis, metoprolol), and they recommeded she follow up with you in a week. I'll follow up and call to schedule an appointment. Good news is she's feeling better now and her heart rate is better (in the 60s). Thank you. | Good morning, I am sorry to hear that this happened, but am glad to hear she is back home. Could you bring her in on [DATE]? [HCW] |
| CLAIR-Short | Question 10: Dear Dr. [HCW], I wanted to let you know that my mother was admitted to [HOSPITAL] on [DATE] for overnight observation, due to having a fast heart rate. She was started on a number of medications (amiodarone, Eliquis, metoprolol), and they recommeded she follow up with you in a | I am glad she is feeling better. I have scheduled her an appt, below. Does this work for you? |

| | | |
|---|---|---|
| | week. I'll follow up and call to schedule an appointment. Good news is she's feeling better now and her heart rate is better (in the 60s). Thank you. | |
| CLAIR-Short | Question 10: Dear Dr. [HCW], I wanted to let you know that my mother was admitted to [HOSPITAL] on [DATE] for overnight observation, due to having a fast heart rate. She was started on a number of medications (amiodarone, Eliquis, metoprolol), and they recommeded she follow up with you in a week. I'll follow up and call to schedule an appointment. Good news is she's feeling better now and her heart rate is better (in the 60s). Thank you. | Thank you for letting me know. I am glad she is feeling better. I will look out for the appointment request to schedule a follow up. [HCW] |
| CLAIR-Short | Question 10: Dear Dr. [HCW], I wanted to let you know that my mother was admitted to [HOSPITAL] on [DATE] for overnight observation, due to having a fast heart rate. She was started on a number of medications (amiodarone, Eliquis, metoprolol), and they recommeded she follow up with you in a week. I'll follow up and call to schedule an appointment. Good news is she's feeling better now and her heart rate is better (in the 60s). Thank you. | Thanks for letting me know. I will be on the lookout for the records. Glad she is feeling better. |
| ChatGPT3 | Question 2: I could really use a sleep aid. Recently I've been having a night or two, sometimes 4, where I just can't sleep. I'm feeling desperate due to lack of sleep and really need something to help me get through this. Is there something you'd recommend? Thanks. | Dear [Patient],<br><br>Thank you for reaching out to me about your recent sleep troubles. I am sorry to hear that you have been experiencing difficulty sleeping and I understand how distressing this can be. I will do my best to provide you with helpful guidance and support.<br><br>There are various reasons why someone may have trouble sleeping, such as stress, anxiety, depression, or physical conditions. Based on what you've shared, it appears that your sleep issues have been ongoing and causing you distress.<br><br>Before considering any sleep aids, I would recommend making some lifestyle changes to improve your sleep hygiene. This includes avoiding caffeine, alcohol, and nicotine before bedtime, establishing a regular sleep schedule, and creating a relaxing bedtime routine. Additionally, make sure your sleeping environment is conducive to sleep, which means keeping it cool, dark, and quiet.<br><br>If these changes do not provide relief, there are non-medical options available, such as cognitive-behavioral therapy for insomnia (CBT-I), which can be effective for treating chronic insomnia. CBT- |

| | | |
|---|---|---|
| | | I focuses on changing negative thought patterns and behaviors that can contribute to insomnia.<br><br>If lifestyle changes and non-medical options are not successful, there are prescription sleep aids available. However, it's important to note that sleep aids should be used as a last resort and only under the guidance of a healthcare professional, due to the potential for side effects and dependency.<br><br>I recommend making an appointment to come in and discuss your options further. During our appointment, we can discuss your medical history and any potential underlying causes for your sleep issues. Together, we can come up with a treatment plan that is tailored to your specific needs and concerns.<br><br>In the meantime, I encourage you to try some of the lifestyle changes I mentioned and to practice relaxation techniques, such as deep breathing and meditation, to help you fall asleep.<br><br>Please don't hesitate to contact me if you have any further questions or concerns. My goal is to help you get the best possible care and support, and I am here to help you through this.<br><br>Best regards,<br><br>[Your Name] |
| ChatGPT4 | Question 2: I could really use a sleep aid. Recently I've been having a night or two, sometimes 4, where I just can't sleep. I'm feeling desperate due to lack of sleep and really need something to help me get through this. Is there something you'd recommend? Thanks. | Dear [Patient's Name],<br><br>I'm sorry to hear that you've been having trouble sleeping recently. It's clear that this is causing you distress and I appreciate you reaching out to discuss it. Lack of sleep can indeed affect our daily activities, mood, and overall health, so addressing this issue is very important.<br><br>While medication can sometimes provide temporary relief, it's typically not the first line of treatment for insomnia or other sleep disturbances. It's important that we first try to identify and address any underlying issues that could be contributing to your sleep problems.<br><br>Are there any changes in your routine or environment that might have coincided with the onset of your sleep problems? Stress, caffeine intake, late meals, or changes in your schedule could all affect your sleep. Making adjustments in these areas may help you sleep better.<br><br>Maintaining a regular sleep schedule, including going to bed and waking up at the same time each day, can also be beneficial. Avoiding screens for an hour before bedtime can also help your body prepare for sleep. This is because screens emit blue light, which can interfere with the production of melatonin, a hormone that regulates sleep.<br><br>In addition to these lifestyle changes, there are relaxation techniques such as deep breathing, progressive muscle relaxation, and mindfulness meditation that can help prepare your body and mind for sleep.<br><br>If these strategies don't provide relief, it would be worth discussing further investigations, such as a |

| | | |
|---|---|---|
| | | sleep study, to rule out conditions like sleep apnea or restless legs syndrome, which could be affecting your sleep.<br><br>As for sleep aids, there are over-the-counter options as well as prescription medications. However, these should be used with caution as they can have side effects and are generally not meant for long-term use. It's best to discuss these options in person, where we can review your medical history and discuss the potential benefits and risks.<br><br>Let's schedule an appointment to discuss this in more detail. I would like to review your symptoms, daily habits, and overall health to help develop a plan that will work best for you. We will work together to improve your sleep quality, and by extension, your overall well-being.<br><br>In the meantime, please keep a sleep diary noting the times you go to bed, fall asleep, wake up, and get out of bed, along with any occurrences that may interrupt your sleep. This can help us identify patterns and potential triggers for your sleep issues.<br><br>Again, I'm really sorry that you're dealing with this, but I'm confident that we can find a solution. Please don't hesitate to reach out if you have any other questions or concerns.<br><br>Sincerely,<br>[Your Name] |
| CLAIR-Short | Question 2: I could really use a sleep aid. Recently I've been having a night or two, sometimes 4, where I just can't sleep. I'm feeling desperate due to lack of sleep and really need something to help me get through this. Is there something you'd recommend? Thanks. | I am sorry to hear that you are not sleeping well. It is important to make sure that you are doing things to help your sleep. Do you have a bedtime routine? Do you limit screen time before bed? Do you have a good sleep environment? Do you try to avoid caffeine after 2 pm? Do you make sure to get regular exercise? I would recommend trying some over the counter melatonin 3 - 5 mg to help you sleep. If this is not helping, we can discuss other options. [HCW] |
| CLAIR-Long | Question 2: I could really use a sleep aid. Recently I've been having a night or two, sometimes 4, where I just can't sleep. I'm feeling desperate due to lack of sleep and really need something to help me get through this. Is there something you'd recommend? Thanks. | Dear Patient,<br><br>Thank you for reaching out to me regarding sleep difficulties that you have been experiencing recently. It sounds like you have been under a lot of stress lately, as it appears that you are having difficulty falling asleep or staying asleep. I am sorry to hear that you are feeling so desperate at times due to your lack of sleep. I understand how frustrating this must be for you, and I hope to provide some information that may be helpful in addressing your concerns.<br><br>First, I would like to gather more information about your current sleep difficulties. How long have you been struggling with these symptoms? Have you noticed any changes in your sleep habits (e.g., going to bed earlier/later than usual, waking up earlier/later than usual)? Are you able to fall asleep easily, but then wake up frequently during the night? Or do you find yourself unable to fall asleep at all? It is also important to ask if you have noticed any other changes in your life recently, such as increased stress levels, changes in diet, or change in medications, as these may affect your ability to sleep. In addition, I would like to know if you have ever experienced similar symptoms in the past. If so, were you able to identify any triggers that caused these episodes? |

| | | |
|---|---|---|
| | | What was most effective in helping you cope with your symptoms at that time?<br><br>Sometimes when we are under a great deal of stress, our bodies respond by entering into what is known as "fight or flight mode." This means that our bodies are preparing themselves to either fight off or flee from a perceived threat. During this time, our bodies release adrenaline, which causes a number of physical changes, including elevated heart rate, rapid breathing, and sweating. These changes make us feel alert and awake, rather than relaxed and ready for sleep. When we are dealing with high levels of anxiety, we may experience episodes of insomnia. In order to decrease the likelihood of experiencing insomnia, it is recommended that we practice good sleep hygiene, as this will create a comfortable and relaxing environment that is conducive to restful sleep. Some tips include avoiding caffeinated drinks after noon, avoiding eating large meals close to bedtime, and limiting screen. |
| Actual Provider Response | Question 2: I could really use a sleep aid. Recently I've been having a night or two, sometimes 4, where I just can't sleep. I'm feeling desperate due to lack of sleep and really need something to help me get through this. Is there something you'd recommend? Thanks. | I would suggest to try melatonin 6 to 9 mg at bedtime to see if that would help you with your sleep. Thank you Dr. [HCW] |
| CLAIR-Short | Question 2: I could really use a sleep aid. Recently I've been having a night or two, sometimes 4, where I just can't sleep. I'm feeling desperate due to lack of sleep and really need something to help me get through this. Is there something you'd recommend? Thanks. | I would be happy to discuss this with you. We can either do a telehealth visit or you can come into the office. I am happy to prescribe a low dose of a sleep aid if you would like. Please let me know and I will get you scheduled. [HCW] |
| CLAIR-Short | Question 2: I could really use a sleep aid. Recently I've been having a night or two, sometimes 4, where I just can't sleep. I'm feeling desperate due to lack of sleep and really need something to help me get through this. Is there something you'd recommend? Thanks. | I want to assure you that it's completely normal to have a night or two where you have trouble falling asleep. However, if you are consistently experiencing trouble sleeping, it's important that you schedule an appointment with me so we can discuss this further. |
| CLAIR-Long | Question 3: Got an upcoming trip to Mexico. Can't control lack of clean water there - might get diarrhea. Could ruin my trip. How about some diarrhea pills for this trip? My friend and their spouse got Rifaximin and Zithromax from their doctor for their trip. | Dear Patient,<br><br>Thank you for reaching out to me regarding your upcoming travel plans to Mexico. I understand how important it is to be healthy during your vacation so that you can enjoy yourself without worrying about falling ill. It sounds like you are concerned about contracting travelers' diarrhea while in Mexico due to unsanitary conditions. This is certainly something to be mindful of as travelers' diarrhea affects 20% to 50% of international travelers. However, I am glad to hear that you are taking precautions by asking ahead of time what medications would be most effective in preventing travelers' diarrhea.<br><br>Travelers' diarrhea is caused by Escherichia coli bacteria (E.coli). These bacteria live in the |

| | | |
|---|---|---|
| | | intestines of humans and animals. While some strains of E.coli are harmless, others produce toxins which cause inflammation of the intestine. The main risk factor for developing travelers' diarrhea is consuming contaminated food or drinking unclean water. In Mexico, it is common for tap water to contain fecal matter which can cause gastrointestinal distress if consumed. Therefore, it is very important to avoid drinking tap water and eating any raw vegetables that may have been washed in it. Other ways to minimize your risk of contracting travelers' diarrhea include boiling all water before consumption, drinking bottled water, avoiding ice cubes made from tap water, washing your hands regularly, and being cautious about eating street food.<br><br>As far as antibiotics go, rifaximin and azithromycin are two commonly prescribed medications used to treat travelers' diarrhea. However, these should only be taken after symptoms develop. If you feel nauseous, vomiting, abdominal cramps, bloody stools, fever, or chills, then you should seek immediate medical attention. Otherwise, you can try using over-the-counter anti-diarrheals such as Pepto Bismol or Imodium to help manage your symptoms.<br><br>I hope this information helps! Please let me know if you have any additional questions. |
| ChatGPT4 | Question 3: Got an upcoming trip to Mexico. Can't control lack of clean water there - might get diarrhea. Could ruin my trip. How about some diarrhea pills for this trip? My friend and their spouse got Rifaximin and Zithromax from their doctor for their trip. | Dear [Patient's Name],<br><br>Thank you for reaching out with your concerns about your upcoming trip to Mexico. Ensuring that you are well-prepared for any potential health concerns while travelling is indeed important.<br><br>It's worth noting that most cases of traveler's diarrhea are mild and resolve on their own within a few days. However, it can sometimes become severe or long-lasting. Preventive measures, such as avoiding tap water, ice, and eating only thoroughly cooked foods, can help reduce your risk.<br><br>Regarding your query about Rifaximin and Zithromax (azithromycin), these antibiotics are sometimes used for the prevention and treatment of traveler's diarrhea, particularly for those who are at a higher risk of complications. However, they are prescription medications and should only be used under the guidance of a healthcare professional.<br><br>Before I can prescribe these medications, I would need to review your medical history and current health status, as these antibiotics might not be suitable for everyone. They can have side effects and can interact with other medications. For instance, Rifaximin is not recommended for people with liver disease and Azithromycin can interact with a variety of other drugs.<br><br>It's also important to note that the overuse of antibiotics can contribute to antibiotic resistance, a significant global health concern. Therefore, it is generally recommended to use these medications for treatment rather than prevention, and only when symptoms are severe or persistent.<br><br>I would suggest that we schedule a pre-travel consultation to discuss this in more detail. During this appointment, we can review your health history, assess your risk, discuss preventive measures, and determine if a prescription for Rifaximin or Zithromax is appropriate for your specific situation. We can also discuss other possible travel-related health issues you might encounter and how to manage them. |

| | | |
|---|---|---|
| | | Meanwhile, please remember to take necessary precautions such as drinking only bottled or boiled water, eating well-cooked food, and frequently washing your hands with soap and water.<br><br>Feel free to message me if you have any more questions or concerns. I hope you have a safe and enjoyable trip!<br><br>Best regards,<br><br>[Your Name, MD] |
| ChatGPT3 | Question 3: Got an upcoming trip to Mexico. Can't control lack of clean water there - might get diarrhea. Could ruin my trip. How about some diarrhea pills for this trip? My friend and their spouse got Rifaximin and Zithromax from their doctor for their trip. | Dear Patient,<br><br>I hope this message finds you well. It's great to hear that you have an upcoming trip to Mexico. I can understand your concern about the availability of clean water there, which might lead to diarrhea and ruin your trip. I will be happy to provide you with some guidance and recommendations.<br><br>Firstly, I would recommend that you take precautions to prevent diarrhea while in Mexico. You can consider drinking only bottled or boiled water, avoiding ice and raw fruits and vegetables, and washing your hands frequently. These measures can go a long way in reducing your risk of getting diarrhea.<br><br>Regarding your request for diarrhea pills, I would like to emphasize that it's important to use antibiotics judiciously and only when necessary. Antibiotics can cause side effects, and their overuse can lead to antibiotic resistance. Therefore, I recommend that you try non-antibiotic medications first, such as loperamide or bismuth subsalicylate, which can be effective in treating diarrhea. I can provide you with a prescription for these medications if you would like.<br><br>However, if your symptoms persist or worsen, or if you develop other symptoms such as fever or severe abdominal pain, it's important that you seek medical attention promptly. In that case, you can contact me, and I will be happy to provide you with further guidance and recommendations.<br><br>I hope that this information is helpful. If you have any further questions or concerns, please don't hesitate to contact me. I wish you a safe and enjoyable trip to Mexico.<br><br>Best regards,<br><br>[Your Name] |
| Actual Provider Response | Question 3: Got an upcoming trip to Mexico. Can't control lack of clean water there - might get diarrhea. Could ruin my trip. How about some diarrhea pills for this trip? My friend and their spouse got Rifaximin and Zithromax from their doctor for their trip. | Hi [PATIENT]! I definitely think you should take antibiotics along on your trip. I prefer azithromycin (Zithromax) - I think it works a little better than rifaxamin. I sent in six 500mg tablets. The course for travelers diarrhea is 3 days, but as you will be in Mexico for a while, I want you to have an additional 3 days if you have diarrhea twice. I put the instructions on the bottle at the pharmacy as well! [HCW] |

| | | |
|---|---|---|
| CLAIR-Short | Question 3: Got an upcoming trip to Mexico. Can't control lack of clean water there - might get diarrhea. Could ruin my trip. How about some diarrhea pills for this trip? My friend and their spouse got Rifaximin and Zithromax from their doctor for their trip. | I sent in Rifaximin to your pharmacy. It is taken for 3 days. You can take it with you and start it if you develop symptoms. |
| CLAIR-Short | Question 3: Got an upcoming trip to Mexico. Can't control lack of clean water there - might get diarrhea. Could ruin my trip. How about some diarrhea pills for this trip? My friend and their spouse got Rifaximin and Zithromax from their doctor for their trip. | I sent in a prescription for you. Have a great trip! |
| CLAIR-Short | Question 3: Got an upcoming trip to Mexico. Can't control lack of clean water there - might get diarrhea. Could ruin my trip. How about some diarrhea pills for this trip? My friend and their spouse got Rifaximin and Zithromax from their doctor for their trip. | [PATIENT], I have sent in a prescription for Cipro to your pharmacy. You should take this medication with you in case you become ill during your trip. It is not intended to take on a regular basis, only if you develop diarrhea. [HCW] |
| ChatGPT3 | Question 4: Hi Doctor [HCW]. I did something to my back this week and I'm having back spasms again. This happens once in a while. Last time, which was a few months ago, I was prescribed cyclobenzaprine 5 mg tablet (FLEXERIL). This really helped me. Can you please renew this prescription and send it to my pharmacy? Thank you! | Dear [Patient],<br><br>Thank you for reaching out to me regarding your back spasms. I am sorry to hear that you are experiencing discomfort again. I understand how frustrating it can be to deal with chronic pain.<br><br>Based on your previous experience with cyclobenzaprine 5 mg tablet (FLEXERIL), it sounds like this medication has been helpful for you in managing your back spasms. However, before renewing your prescription, I would like to schedule an appointment with you to discuss your symptoms and review your medical history.<br><br>During this appointment, we can also discuss alternative treatment options and any potential side effects or interactions with other medications you may be taking. It's important to ensure that the medication is still the best course of action for your condition, and that there are no changes to your overall health that may impact its effectiveness or safety.<br><br>Please let me know your availability, and I will be happy to schedule an appointment for you at the earliest convenience. In the meantime, I recommend that you try applying heat or ice to the affected area, and gentle stretches to help alleviate your discomfort.<br><br>Thank you for trusting me with your care. I look forward to seeing you soon.<br><br>Best regards,<br>[HCW] |
| ChatGPT4 | Question 4: Hi Doctor [HCW]. I did something to my back this week and I'm having back spasms again. This happens | Dear [Patient's Name],<br><br>I'm sorry to hear about your back spasms. These can indeed be very painful and disruptive. I |

| | | |
|---|---|---|
| | once in a while. Last time, which was a few months ago, I was prescribed cyclobenzaprine 5 mg tablet (FLEXERIL). This really helped me. Can you please renew this prescription and send it to my pharmacy? Thank you! | appreciate your reaching out to me so we can work together to help alleviate your discomfort.<br><br>Your previous experience with cyclobenzaprine (Flexeril) seems to have provided good relief, and I'm glad to hear it was effective for you. Given your past positive response, it's certainly worth considering again.<br><br>However, before renewing the prescription, it would be best for us to have a quick check-in, either via telehealth or in person. This is to ensure there are no new medical issues or contraindications that we should be aware of, and to discuss any recent changes in your health status or medications. It's always essential to do this before prescribing muscle relaxants, as these can sometimes interact with other medications or conditions.<br><br>While we manage the immediate issue, we also need to think about the bigger picture. Since this is a recurring issue, I would like to explore preventative strategies with you to help decrease the frequency and severity of these spasms. We can discuss potential causes, like posture or lifting techniques, and possible interventions, such as physical therapy, exercises, or lifestyle modifications, to prevent this from happening in the future.<br><br>In the meantime, here are a few suggestions to help alleviate your discomfort:<br>1. Try to rest your back. Avoid heavy lifting and any activities that seem to worsen your back pain.<br>2. Apply a warm compress or a heating pad on a low setting to the affected area.<br>3. Over-the-counter pain relievers, such as ibuprofen, could be used in the short term, unless there's a reason you can't take these medications.<br><br>Please arrange a time for a consultation at your earliest convenience so we can further discuss your symptoms and treatment plan. Remember, your health and comfort are my top priority.<br><br>Thank you for reaching out and I look forward to speaking with you soon.<br><br>Best regards,<br>Dr. [HCW] |
| CLAIR-Long | Question 4: Hi Doctor [HCW]. I did something to my back this week and I'm having back spasms again. This happens once in a while. Last time, which was a few months ago, I was prescribed cyclobenzaprine 5 mg tablet (FLEXERIL). This really helped me. Can you please renew this prescription and send it to my pharmacy? Thank you! | Dear Patient,<br><br>I am sorry to hear about your back spasm issue. I understand how uncomfortable and frustrating this problem can be. Let me share some information on back spasms with you. Back spasms are involuntary contractions of muscles causing pain and discomfort. Spasms often occur when muscles become strained or injured as a result of overuse or improper lifting. They can also result from a pinched nerve or arthritis. Lumbar spinal stenosis is another common cause of lower back spasms.<br><br>I would recommend applying heat to the affected area, using ice packs, taking a warm bath, and gentle stretching exercises such as yoga to relieve symptoms. Additionally, if you are experiencing pain due to inflammation, you may want to try anti-inflammatory medications like ibuprofen or naproxen. Furthermore, you should avoid sleeping on your stomach, if possible. You can also use pillows underneath your knees to reduce stress on your lower back. If these methods do not work, |

| | | |
|---|---|---|
| | | I would suggest making an appointment to see one of our providers here at [HOSPITAL] so we can evaluate the source of your back spasm and come up with a treatment plan that will help you find relief.<br><br>Thank you for reaching out to us, and I hope you feel better soon. |
| Actual Provider Response | Question 4: Hi Doctor [HCW]. I did something to my back this week and I'm having back spasms again. This happens once in a while. Last time, which was a few months ago, I was prescribed cyclobenzaprine 5 mg tablet (FLEXERIL). This really helped me. Can you please renew this prescription and send it to my pharmacy? Thank you! | Refill for flexeril sent to your pharmacy. If back pain is severe, not improving, or associated with new leg weakness please let us know. When taking flexeril, avoid taking it while driving. It can make you very drowsy.[HCW] |
| CLAIR-Short | Question 4: Hi Doctor [HCW]. I did something to my back this week and I'm having back spasms again. This happens once in a while. Last time, which was a few months ago, I was prescribed cyclobenzaprine 5 mg tablet (FLEXERIL). This really helped me. Can you please renew this prescription and send it to my pharmacy? Thank you! | [PATIENT], I am sorry to hear that you are having back spasms. I see that [HCW] prescribed this for you last time. Have you been taking any over - the - counter medications? Any relief? |
| CLAIR-Short | Question 4: Hi Doctor [HCW]. I did something to my back this week and I'm having back spasms again. This happens once in a while. Last time, which was a few months ago, I was prescribed cyclobenzaprine 5 mg tablet (FLEXERIL). This really helped me. Can you please renew this prescription and send it to my pharmacy? Thank you! | I am sorry to hear that you are having back spasms. Since you are having back spasms, I will send in a muscle relaxer for you. I would also recommend that you follow up with Ortho Spine. I will place the referral. Please let me know if you have any further questions. Thanks, [HCW] |
| CLAIR-Short | Question 4: Hi Doctor [HCW]. I did something to my back this week and I'm having back spasms again. This happens once in a while. Last time, which was a few months ago, I was prescribed cyclobenzaprine 5 mg tablet (FLEXERIL). This really helped me. Can you please renew this prescription and send it to my pharmacy? Thank you! | I sent in a refill of the cyclobenzaprine. Hope you feel better soon! |
| ChatGPT4 | Question 5: Hello doctor. I've had a nonproductive dry cough for about 3 weeks. I've tried cough syrup and cough drops, but | Dear [PATIENT],<br><br>Thank you for reaching out and I'm sorry to hear that you're not feeling well. The information you |

| | | |
|---|---|---|
| | nothing seems to help and it's keeping me up at night. My sister mentioned she had something similar happen with a dry hacking cough when she took lisinopril, and her doctor said it was a side effect. I noticed the cough and the tickle in my throat after we last increased the dose of lisinopril. Could I be having a side effect too? Thanks, [PATIENT] | provided is indeed very helpful.<br><br>Based on your symptoms and the timing of their onset, it's possible that the increase in your lisinopril dosage may be contributing to your persistent dry cough. Lisinopril, which is an angiotensin-converting enzyme (ACE) inhibitor, can cause a dry cough in some patients. This side effect is not harmful, but can be quite bothersome, especially if it's affecting your sleep.<br><br>Before making any conclusions, it's important that we rule out other potential causes of your symptoms. These could include allergies, respiratory infections, or gastroesophageal reflux disease (GERD), among others.<br><br>I would recommend that we schedule an appointment to discuss this in more detail. In the meantime, I advise you to keep track of any additional symptoms you may be experiencing. Please do not stop or adjust the dosage of your medication without consulting with me first, as it's crucial for managing your blood pressure.<br><br>During our upcoming appointment, we can consider alternatives to lisinopril, if indeed it is causing your cough. There are other types of blood pressure medications available that might be better suited to you, such as angiotensin receptor blockers (ARBs), which tend to cause fewer side effects.<br><br>Please remember that this message is meant to provide general advice and does not replace a formal evaluation. If your symptoms worsen or if you develop additional symptoms such as fever, difficulty breathing, or chest pain, please seek immediate medical attention.<br><br>I appreciate your proactive approach to your health and look forward to our discussion.<br><br>Best regards,<br>[Doctor] |
| ChatGPT3 | Question 5: Hello doctor. I've had a nonproductive dry cough for about 3 weeks. I've tried cough syrup and cough drops, but nothing seems to help and it's keeping me up at night. My sister mentioned she had something similar happen with a dry hacking cough when she took lisinopril, and her doctor said it was a side effect. I noticed the cough and the tickle in my throat after we last increased the dose of lisinopril. Could I be having a side effect too? Thanks, [PATIENT] | Dear [PATIENT],<br><br>Thank you for reaching out to me regarding your persistent dry cough. I understand how frustrating and disruptive it can be to experience symptoms that interfere with your daily life, especially when you have tried over-the-counter remedies without relief.<br><br>Based on what you've described, it is possible that your cough may be related to a side effect of lisinopril. As you may know, lisinopril is a medication commonly used to treat high blood pressure and heart failure, but it can cause coughing in some patients as a side effect.<br><br>Given the timing of your symptoms starting after an increase in the dose of lisinopril, it is important to explore this possibility further. I would like to invite you to schedule an appointment with me as soon as possible so that we can evaluate your condition and discuss potential next steps.<br><br>During the appointment, I will ask you some questions about your medical history and perform a physical examination to help me understand your symptoms better. Depending on our findings, we |

| | | |
|---|---|---|
| | | may consider adjusting your medication, changing to a different medication, or exploring other possible causes of your cough.<br><br>In the meantime, I recommend that you continue to use cough drops or honey to help soothe your throat and alleviate your symptoms. It is also essential to stay well-hydrated and avoid exposure to irritants such as smoke, dust, or other environmental triggers that can exacerbate your cough.<br><br>I hope this information helps, and I look forward to seeing you soon. Please feel free to contact me if you have any further questions or concerns.<br><br>Take care and stay healthy.<br><br>Best regards,<br>[DOCTOR] |
| CLAIR-Long | Question 5: Hello doctor. I've had a nonproductive dry cough for about 3 weeks. I've tried cough syrup and cough drops, but nothing seems to help and it's keeping me up at night. My sister mentioned she had something similar happen with a dry hacking cough when she took lisinopril, and her doctor said it was a side effect. I noticed the cough and the tickle in my throat after we last increased the dose of lisinopril. Could I be having a side effect too? Thanks, [PATIENT] | Dear Patient,<br><br>Thank you for reaching out to us regarding your recent symptoms. It sounds like you've been experiencing a persistent cough over the past few weeks, which has kept you awake at night. You mention that you started taking Lisinopril recently as well, and wonder whether these two could be related.<br><br>Firstly, it is important to note that many factors can lead to a persistent cough, including viral upper respiratory tract infections (such as colds), allergies, gastroesophageal reflux disease (GERD) and postnasal drip. In addition, certain medications such as angiotensin converting enzyme inhibitors (ACEIs) and angiotensin II receptor blockers (ARBs) can cause a chronic dry cough in some individuals. If you believe that this may be the case, I would encourage you to reach out to the prescribing physician to discuss alternative options. It is also important to note that although ACEIs and ARBs may be associated with a cough, they are often very effective treatments for high blood pressure.<br><br>Without knowing more about your medical history, it is difficult to assess what might be causing your cough. However, given that you have been experiencing it for several weeks now, it is likely that an underlying condition is responsible. For this reason, I strongly recommend that you schedule an appointment with one of our physicians so that you receive the appropriate treatment.<br><br>If you find yourself struggling to breathe during your episodes of coughing, or if you develop additional symptoms such as fever, chest pain, shortness of breath, difficulty swallowing, or loss of consciousness, please seek emergent care. |
| CLAIR-Short | Question 5: Hello doctor. I've had a nonproductive dry cough for about 3 weeks. I've tried cough syrup and cough drops, but nothing seems to help and it's keeping me up at night. My sister mentioned she had something similar happen with a dry hacking cough when she took lisinopril, and her | I am sorry to hear that you are having a cough. I agree with you that it would be reasonable to try stopping the lisinopril for a few days to see if the cough improves. If it does, then we can make a plan for a different medication. If it does not, then we will need to investigate further. Please let me know. [HCW] |

| | | |
|---|---|---|
| | doctor said it was a side effect. I noticed the cough and the tickle in my throat after we last increased the dose of lisinopril. Could I be having a side effect too? Thanks, [PATIENT] | |
| Actual Provider Response | Question 5: Hello doctor. I've had a nonproductive dry cough for about 3 weeks. I've tried cough syrup and cough drops, but nothing seems to help and it's keeping me up at night. My sister mentioned she had something similar happen with a dry hacking cough when she took lisinopril, and her doctor said it was a side effect. I noticed the cough and the tickle in my throat after we last increased the dose of lisinopril. Could I be having a side effect too? Thanks, [PATIENT] | [PATIENT], [HCW] reviewed your message and would like you to stop the Lisinopril, she sent in Losartan 50mg to take daily. The cough should improve over 2 weeks. Let us know if you have further questions/concerns. Thank you. |
| CLAIR-Short | Question 5: Hello doctor. I've had a nonproductive dry cough for about 3 weeks. I've tried cough syrup and cough drops, but nothing seems to help and it's keeping me up at night. My sister mentioned she had something similar happen with a dry hacking cough when she took lisinopril, and her doctor said it was a side effect. I noticed the cough and the tickle in my throat after we last increased the dose of lisinopril. Could I be having a side effect too? Thanks, [PATIENT] | [PATIENT], It is possible that this is a side effect of the lisinopril. I have sent in a prescription for a cough syrup to help you get some rest at night. I would like you to stop the lisinopril for 1 week and see if your cough improves. If it does, we will know the lisinopril was the cause. If it doesn't, we will need to look at other causes. Please let me know how you are doing and keep me updated. [HCW] |
| CLAIR-Short | Question 5: Hello doctor. I've had a nonproductive dry cough for about 3 weeks. I've tried cough syrup and cough drops, but nothing seems to help and it's keeping me up at night. My sister mentioned she had something similar happen with a dry hacking cough when she took lisinopril, and her doctor said it was a side effect. I noticed the cough and the tickle in my throat after we last increased the dose of lisinopril. Could I be having a side effect too? Thanks, [PATIENT] | Thank you for reaching out. I am sorry to hear you have been experiencing these symptoms. I would recommend having you be evaluated in our clinic or a Walk - In Clinic to be evaluated for your symptoms. If you are not able to be evaluated in the next 24 - 48 hours, I would recommend going to an ER. I hope you feel better soon! - [HCW] |
| ChatGPT4 | Question 6: Good Afternoon. I recently had some genetic testing performed, since I am trying to conceive with my partner. My | Dear [Patient],<br><br>Thank you for reaching out and for being proactive about your health, especially in regard to your |

| | | |
|---|---|---|
| | results showed that I'm a carrier for hemochromatosis. The fertility clinic recommended I reach out to you about these results. Is there anything I need to do? Thanks in advance, [Patient] | plans of conceiving a child. Genetic testing is a valuable tool that can give us insight into potential health risks, and in your case, it has identified you as a carrier for hemochromatosis.<br><br>Hemochromatosis is a genetic condition that causes the body to absorb too much iron from the diet. This excess iron is then stored in the body's organs, particularly the liver, heart, and pancreas, which can lead to life-threatening conditions such as liver disease, heart problems, and diabetes.<br><br>As a carrier, you have one copy of the gene mutation but not the disease itself. This generally means you are unlikely to develop hemochromatosis or show symptoms. However, the implications of this result may be different for your future child, depending on whether your partner is also a carrier.<br><br>If both parents are carriers, there is a 25% chance with each pregnancy that the child could inherit two copies of the gene mutation, one from each parent, and therefore develop the disease. Alternatively, there is a 50% chance the child could be a carrier like you, and a 25% chance the child will neither be a carrier nor have the disease.<br><br>To better understand this risk, it would be beneficial for your partner to also have genetic testing performed. If your partner is found to be a carrier, you may wish to consult with a genetic counselor who can provide more detailed information and discuss possible options with you.<br><br>In terms of your personal health, being a carrier for hemochromatosis doesn't usually require specific management, but routine check-ups and maintaining a healthy lifestyle are always recommended. However, in the context of your fertility journey, it would be wise to avoid iron supplements unless specifically recommended by your healthcare provider.<br><br>Thank you again for your proactive approach to your health. Please let me know if your partner decides to undergo genetic testing or if you have any further questions or concerns. Remember, I'm here to support you in your journey to parenthood and overall well-being.<br><br>Best regards,<br><br>[Your Name] |
| ChatGPT3 | Question 6: Good Afternoon. I recently had some genetic testing performed, since I am trying to conceive with my partner. My results showed that I'm a carrier for hemochromatosis. The fertility clinic recommended I reach out to you about these results. Is there anything I need to do? Thanks in advance, [Patient] | Dear [Patient],<br><br>Good afternoon, thank you for reaching out to me about the results of your recent genetic testing. I understand that you and your partner are trying to conceive, and it is important to ensure that you receive the best possible care and support during this process.<br><br>I appreciate you sharing that you are a carrier for hemochromatosis. This is a genetic condition that causes excess iron to build up in the body, which can lead to serious health problems if left untreated. It is important to note that being a carrier means that you have one copy of the gene mutation that causes hemochromatosis, but it does not mean that you have the condition itself.<br><br>Given your situation, I recommend that you schedule an appointment with me to discuss your |

| | | |
|---|---|---|
| | | results in more detail. During this appointment, we can discuss your specific situation and any potential implications for your health and the health of your future child. It is important that we develop a plan to manage this condition in the context of your plans for conception.<br><br>As part of our discussion, we can explore various options for managing hemochromatosis, including genetic counseling, monitoring iron levels, and potentially starting treatment if necessary. I will also be able to answer any additional questions you may have and provide guidance on next steps.<br><br>Thank you again for reaching out to me, [Patient]. I am committed to providing you with the best possible care and support, and I look forward to discussing your situation in more detail during our upcoming appointment.<br><br>Sincerely,<br>[Your Name] |
| CLAIR-Long | Question 6: Good Afternoon. I recently had some genetic testing performed, since I am trying to conceive with my partner. My results showed that I'm a carrier for hemochromatosis. The fertility clinic recommended I reach out to you about these results. Is there anything I need to do? Thanks in advance, [Patient] | Dear Patient,<br><br>Thank you for reaching out to me regarding your recent genetic test results. It sounds like you were tested for hereditary hemochromatosis (HH), which is a condition where your body absorbs too much iron from food. When excess iron builds up in your body, it can be harmful to many organs such as the liver, heart, pancreas, joints, and skin. HH is caused by a mutation in a gene called HFE. This gene produces protein that helps your body regulate how much iron you absorb from your diet. You may have inherited one copy of a mutated HFE gene from each parent, which means you would be a "carrier" for HH. However, if only one of your parents has a mutated HFE gene, then you will have HH.<br><br>It is important to note that being a carrier does not mean you will develop symptoms or complications of HH. In fact, most people who carry a mutated HFE gene never develop any symptoms. However, if you become pregnant, you should discuss with your OBGYN whether or not you should be screened for HH during your pregnancy. If you do have HH and become pregnant, your healthcare provider can monitor your iron levels to make sure they stay stable.<br><br>If you have additional questions or concerns, please let me know. Otherwise, I hope this information was helpful! Take care and have a great weekend. |
| CLAIR-Short | Question 6: Good Afternoon. I recently had some genetic testing performed, since I am trying to conceive with my partner. My results showed that I'm a carrier for hemochromatosis. The fertility clinic recommended I reach out to you about these results. Is there anything I need to do? Thanks in advance, [Patient] | Can you please send us a copy of the test results? |

| | | |
|---|---|---|
| Actual Provider Response | Question 6: Good Afternoon. I recently had some genetic testing performed, since I am trying to conceive with my partner. My results showed that I'm a carrier for hemochromatosis. The fertility clinic recommended I reach out to you about these results. Is there anything I need to do? Thanks in advance, [Patient] | Hi, [PATIENT]! Thanks for letting me know! Fortunately, your most recent liver labs look good. Hemochromatosis is a disease where you absorb too much iron due to a genetic defect and the iron gets stored in your organs. We can monitor it over time. Sometimes, people are treated later in life with intermittent phlebotomy (removing blood to take away excess iron). I will send labs to check your iron levels and see how things are doing for now. Please run by the lab at your convenience, and I will follow up! Here is a nice, reputable summary of HH: cdc.gov/genomics/disease/hemochromatosis.htm#:~:text=Hereditary%20hemochromatosis%20is %20a%20genetic, about%20testing%20for%20hereditary%20hemochromatosis. We can also talk at our next clinic visit in more detail. [HCW] |
| CLAIR-Short | Question 6: Good Afternoon. I recently had some genetic testing performed, since I am trying to conceive with my partner. My results showed that I'm a carrier for hemochromatosis. The fertility clinic recommended I reach out to you about these results. Is there anything I need to do? Thanks in advance, [Patient] | I'm going to send you to a hematologist to discuss this further. I have placed the referral. [HCW] |
| CLAIR-Short | Question 6: Good Afternoon. I recently had some genetic testing performed, since I am trying to conceive with my partner. My results showed that I'm a carrier for hemochromatosis. The fertility clinic recommended I reach out to you about these results. Is there anything I need to do? Thanks in advance, [Patient] | Thanks for the message. I reviewed the testing. It is a common variant that is not associated with disease. You don't have to do anything special. |
| CLAIR-Long | Question 7: Dear Doctor [HCW], I had two weeks of a bad cold. Never had a fever, and I tested negative for covid, but my cough won't go away even a couple weeks later, and my energy level isn't great. I'm having to take an allergy pill every day just to go to sleep. Do I need a flu test, or an allergy test? | Dear Patient,<br><br>Thank you for reaching out to me regarding your lingering cough after having a bad cold. I understand how frustrating it can be when you think you are over an illness only to find yourself still feeling unwell. It sounds like you may have developed a postinfectious cough that has been persisting since your cold and causing you to feel fatigued. This type of cough can last up to eight weeks after the initial upper respiratory tract infection. There are several treatment options available to help reduce the duration and severity of symptoms associated with this type of cough.<br><br>I would recommend using a humidifier at night to help loosen mucus in your airways, which will allow you to cough it up more easily. If you do not already own one, you can purchase one online or at any major pharmacy. Another option is to use a saline nasal spray once daily to help thin mucus in your nose and sinuses. You can also try drinking warm liquids such as tea with honey to help soothe your throat and loosen congestion. Gargling with warm salt water a few times per day can also help alleviate some of your symptoms. Over-the-counter medications such as Mucinex can be used to thin secretions in your lungs to make them easier to expel through coughing. As always, please consult with your pharmacist before taking any new medication to ensure there are no interactions with other medications you are currently taking. |

| | | |
|---|---|---|
| | | I hope this information helps you manage your symptoms. Please let me know if you have any questions or concerns.<br><br>Best, |
| ChatGPT3 | Question 7: Dear Doctor [HCW], I had two weeks of a bad cold. Never had a fever, and I tested negative for covid, but my cough won't go away even a couple weeks later, and my energy level isn't great. I'm having to take an allergy pill every day just to go to sleep. Do I need a flu test, or an allergy test? | Dear [Patient],<br><br>Thank you for reaching out to me regarding your ongoing symptoms. I'm sorry to hear that you have been experiencing a persistent cough and low energy levels, despite testing negative for COVID-19. I understand that this can be frustrating and disruptive to your daily life.<br><br>Based on your description of your symptoms, it is possible that your cold may have progressed into a secondary bacterial infection, such as bronchitis. Alternatively, you may be experiencing allergies that are exacerbating your symptoms.<br><br>Before considering any tests, I would like to conduct a physical examination to better understand the nature of your symptoms. During the examination, I will listen to your lungs and check for any signs of congestion or infection. Based on my findings, we can determine if additional tests or treatment are necessary.<br><br>In the meantime, I recommend that you continue taking your allergy medication as directed. Additionally, you can try using a humidifier in your bedroom at night to help alleviate any congestion or dryness in your nasal passages.<br><br>If your symptoms persist or worsen, please do not hesitate to reach out to me. I am here to provide you with the best possible care and support. Let's work together to get you feeling better soon.<br><br>Best regards,<br><br>[Your name] |
| ChatGPT4 | Question 7: Dear Doctor [HCW], I had two weeks of a bad cold. Never had a fever, and I tested negative for covid, but my cough won't go away even a couple weeks later, and my energy level isn't great. I'm having to take an allergy pill every day just to go to sleep. Do I need a flu test, or an allergy test? | Dear [Patient's Name],<br><br>Thank you for reaching out and I'm sorry to hear that you've been feeling unwell. It's important to manage and understand your symptoms, so your diligence in keeping track of them is appreciated.<br><br>Cold symptoms can often linger, especially the cough, which can persist for several weeks after the other symptoms have resolved. This is known as a post-viral cough, and it's often due to the inflammation in your airways caused by the viral infection. However, it's concerning that your energy level has not improved.<br><br>The use of an allergy pill suggests that you might be dealing with some sort of allergic reaction, possibly even to something in your environment. If the allergy pill is helping you sleep but not addressing your cough or energy level, it would be beneficial to further investigate the root cause of these symptoms.<br><br>At this point, a flu test is not necessary, as your symptoms don't match the common symptoms of |

| | | |
|---|---|---|
| | | influenza, like fever, chills, body aches, etc. However, it might be worthwhile to explore an allergy test to rule out any potential allergens that might be causing your symptoms. This would especially be the case if your symptoms persist or worsen.<br><br>Another possible cause could be a condition known as post-viral fatigue syndrome, which can occur after a viral illness, and may explain your low energy levels. This condition often resolves over time, but it's important to manage it properly to avoid prolonging the recovery process.<br><br>To help you get a more precise diagnosis and treatment plan, I would suggest scheduling an appointment so we can discuss your symptoms in detail. It would be helpful if you can take note of any potential triggers or changes in your environment, diet, or lifestyle that may have coincided with the onset of your symptoms.<br><br>In the meantime, make sure to stay hydrated, eat nutritious foods, get plenty of rest, and avoid any known triggers of your symptoms. These measures should help support your immune system as it recovers.<br><br>Remember, I'm here to support you through this process. Your health and well-being are my top priority. Please feel free to reach out if you have any other concerns or questions.<br><br>Best,<br><br>Dr. [HCW] |
| CLAIR-Short | Question 7: Dear Doctor [HCW], I had two weeks of a bad cold. Never had a fever, and I tested negative for covid, but my cough won't go away even a couple weeks later, and my energy level isn't great. I'm having to take an allergy pill every day just to go to sleep. Do I need a flu test, or an allergy test? | I'm sorry to hear you're not feeling well. I would recommend you be seen in a walk in clinic for evaluation. We don't have any openings in our clinic today. They can test you for flu and strep if needed. Thank you, [HCW] |
| Actual Provider Response | Question 7: Dear Doctor [HCW], I had two weeks of a bad cold. Never had a fever, and I tested negative for covid, but my cough won't go away even a couple weeks later, and my energy level isn't great. I'm having to take an allergy pill every day just to go to sleep. Do I need a flu test, or an allergy test? | Ok, Dr. [HCW] asks do you have other symptoms or is it just a lingering cough? That's a typical pattern after a respiratory infection because the airways are still irritated. The cough can linger for several weeks. No testing needed right now. Would you like us to send you in some tessalon perles to help your cough? If so, which pharmacy do you prefer? Thank you, [HCW] |
| CLAIR-Short | Question 7: Dear Doctor [HCW], I had two weeks of a bad cold. Never had a fever, and I tested negative for covid, but my cough won't go away even a couple weeks later, and my energy level isn't great. I'm having to | Are you having any other symptoms? Any shortness of breath? Any sinus congestion? |

| | | |
|---|---|---|
| | take an allergy pill every day just to go to sleep. Do I need a flu test, or an allergy test? | |
| CLAIR-Short | Question 7: Dear Doctor [HCW], I had two weeks of a bad cold. Never had a fever, and I tested negative for covid, but my cough won't go away even a couple weeks later, and my energy level isn't great. I'm having to take an allergy pill every day just to go to sleep. Do I need a flu test, or an allergy test? | Have you tried any over the counter cough medicines? Have you tried mucinex? Any shortness of breath or chest pain? Any dizziness or weakness? Thanks |
| ChatGPT4 | Question 8: I'm currently pregnant and have been having an issue with passing bright red blood with my bowel movements over the past year. At first it was infrequent, but for the past week has been almost daily. Every time I pass stool, there's bright red blood, as well as some abdominal pain which goes away after the BM. I'm guessing they may not do colonscopies during pregnancy, but I was thinking I should get this checked out. Please let me know any advice you have? Thanks, [PATIENT] | Dear [PATIENT],<br><br>I appreciate your message and your proactive approach to your health, particularly during your pregnancy. I want to assure you that we take your concerns very seriously.<br><br>The symptoms you're experiencing, notably the bright red blood in your stool and abdominal pain, are indeed concerning and warrant prompt medical attention. The bright red color often indicates bleeding in the lower part of the digestive tract, such as the rectum or colon. This could be due to various conditions like hemorrhoids, anal fissures, or in rare cases, more serious conditions like colorectal polyps or cancer. Given your ongoing pregnancy and the recent increase in frequency, we need to proceed with caution and diligence.<br><br>You're correct that colonoscopies are typically avoided during pregnancy unless absolutely necessary due to the risk to both the mother and the unborn child. However, there are other less invasive procedures that we can use to understand the cause of this bleeding, such as a flexible sigmoidoscopy or an anoscopy, which can be safely done during pregnancy.<br><br>I would advise scheduling an appointment with me as soon as possible so we can further discuss these symptoms and decide on the best next steps for you and your baby's health. We will likely refer you to a gastroenterologist who specializes in these types of conditions and is experienced in treating pregnant women. In the meantime, please keep note of any changes or additional symptoms you may experience.<br><br>Your comfort and well-being, as well as that of your unborn child, are our utmost priority. Please don't hesitate to reach out if you have any further questions or concerns. Thank you for your trust in your care.<br><br>Best,<br>[Your Name] |
| ChatGPT3 | Question 8: I'm currently pregnant and have been having an issue with passing bright red blood with my bowel movements over the past year. At first it was infrequent, but for | Dear [Patient],<br><br>Thank you for reaching out to me regarding your current issue. I'm sorry to hear that you've been experiencing bright red blood with your bowel movements for the past year, which has now |

| | | |
|---|---|---|
| | the past week has been almost daily. Every time I pass stool, there's bright red blood, as well as some abdominal pain which goes away after the BM. I'm guessing they may not do colonscopies during pregnancy, but I was thinking I should get this checked out. Please let me know any advice you have? Thanks, [PATIENT] | become almost a daily occurrence, along with abdominal pain that subsides after a bowel movement. I understand that you're currently pregnant, and you're wondering if there are any restrictions on diagnostic testing that you should be aware of.<br><br>Based on your symptoms, it's important to get this evaluated to identify the underlying cause of your symptoms. While colonoscopies may not be typically recommended during pregnancy, there are other diagnostic tests that can be done to investigate the source of the bleeding. Some of these tests include flexible sigmoidoscopy, stool tests, and blood tests. I can refer you to an OB-GYN specialist or a gastroenterologist who is trained in diagnosing and treating pregnant women.<br><br>It's important to note that bright red blood with bowel movements can indicate various underlying conditions such as hemorrhoids, anal fissures, or inflammatory bowel disease (IBD). Hemorrhoids are the most common cause of bleeding during bowel movements, especially during pregnancy, but it's essential to rule out other potential causes as well. An accurate diagnosis is critical to ensure the best possible outcome for you and your baby.<br><br>I recommend that you schedule an appointment to see a specialist as soon as possible to determine the cause of your symptoms. Additionally, I suggest keeping a record of your symptoms, such as when the bleeding and abdominal pain occur, and if there are any other symptoms or factors that aggravate them. It can help the specialist diagnose the problem accurately.<br><br>I understand that this situation may cause you some worry, but please be assured that I'm here to support you and will provide any guidance and assistance that you need throughout this process. Please let me know if you have any further questions or concerns.<br><br>Best regards,<br>[Your Name] |
| CLAIR-Long | Question 8: I'm currently pregnant and have been having an issue with passing bright red blood with my bowel movements over the past year. At first it was infrequent, but for the past week has been almost daily. Every time I pass stool, there's bright red blood, as well as some abdominal pain which goes away after the BM. I'm guessing they may not do colonscopies during pregnancy, but I was thinking I should get this checked out. Please let me know any advice you have? Thanks, [PATIENT] | Dear Patient,<br><br>Thank you for reaching out to me regarding your concerns about stools containing bright red blood. I understand how worried you must be feeling right now, especially given your current pregnancy status. However, I hope to alleviate some of these fears by discussing what might be causing your symptoms and when you should seek medical attention.<br><br>The most common cause of rectal bleeding in women who are pregnant is hemorrhoids. Hemorrhoids occur due to increased pressure on the veins of the rectum and can be exacerbated by constipation. This is typically associated with painless bleeding, although sometimes there is a small amount of discomfort. The good news is that hemorrhoids tend to resolve themselves after delivery. If you are concerned about your hemorrhoids, please consider taking sitz baths and applying topical medications such as Preparation H to help improve your symptoms.<br><br>Another common cause of bleeding is anal fissures, which are small tears in the skin around the anus. These often result from straining to pass a hard stool. In addition to bloody stools, you might experience painful bowel movements and a burning sensation around the anus. You can try using stool softeners to reduce the risk of tearing and also apply topical creams to relieve discomfort. |

| | | |
|---|---|---|
| | | If you are experiencing severe pain or heavy bleeding, including clots or black tarry stools, then you need to see your obstetrician immediately. It is important to rule out other conditions, such as colon cancer or diverticulitis, which can lead to serious complications. Additionally, if you have recently had any vaginal bleeding or spotting, then you should contact your OBGYN. |
| CLAIR-Short | Question 8: I'm currently pregnant and have been having an issue with passing bright red blood with my bowel movements over the past year. At first it was infrequent, but for the past week has been almost daily. Every time I pass stool, there's bright red blood, as well as some abdominal pain which goes away after the BM. I'm guessing they may not do colonscopies during pregnancy, but I was thinking I should get this checked out. Please let me know any advice you have? Thanks, [PATIENT] | I'm sorry to hear this! Have you had any recent trauma to the area or have you been constipated? Is the pain on the right or left side? Is it sharp or dull? Any nausea or vomiting? When was your last BM? [HCW] |
| CLAIR-Short | Question 8: I'm currently pregnant and have been having an issue with passing bright red blood with my bowel movements over the past year. At first it was infrequent, but for the past week has been almost daily. Every time I pass stool, there's bright red blood, as well as some abdominal pain which goes away after the BM. I'm guessing they may not do colonscopies during pregnancy, but I was thinking I should get this checked out. Please let me know any advice you have? Thanks, [PATIENT] | I am sorry to hear this. I would like to refer you to a GI doctor to get this checked out. I have placed the referral and they should be contacting you soon to schedule an appointment. [HCW] |
| Actual Provider Response | Question 8: I'm currently pregnant and have been having an issue with passing bright red blood with my bowel movements over the past year. At first it was infrequent, but for the past week has been almost daily. Every time I pass stool, there's bright red blood, as well as some abdominal pain which goes away after the BM. I'm guessing they may not do colonscopies during pregnancy, but I was thinking I should get this checked out. Please let me know any advice you have? Thanks, [PATIENT] | Hi [PATIENT]. Given your symptoms, I would absolutely recommend a check in with GI. I am not sure whether they would proceed with colonoscopy, but it is worth a discussion with the provider. I would be happy to initiate a referral for you--would you like for me to pull that trigger? Thank you for reaching out, Dr. [HCW] |
| CLAIR-Short | Question 8: I'm currently pregnant and have been having an issue with passing bright red blood with my bowel movements over the | I would advise you to go to the ER to receive medical attention. |

| | | |
|---|---|---|
| | past year. At first it was infrequent, but for the past week has been almost daily. Every time I pass stool, there's bright red blood, as well as some abdominal pain which goes away after the BM. I'm guessing they may not do colonscopies during pregnancy, but I was thinking I should get this checked out. Please let me know any advice you have? Thanks, [PATIENT] | |
| ChatGPT3 | Question 9: Hello, I wanted to let you know I tested positive for covid today. I'm having a cough, dry throat, feeling tired, and a small headache. No fevers or aches. I'm up-to-date with my covid vaccine so I'm hoping things won't get worse. I'm trying to rest, doing some nasal rinses, and steam. Anything else specific you'd recommend me to do to treat? Thanks. [PATIENT] | Dear [Patient],<br><br>I am sorry to hear that you tested positive for COVID-19. I hope you are feeling okay despite the symptoms you have described, and I am here to provide you with guidance on what you can do to manage them. Thank you for reaching out to me.<br><br>It is great to hear that you have already started taking some self-care measures by getting rest, doing nasal rinses, and steam. These are excellent ways to ease the symptoms that you are experiencing. In addition to these, there are some other things that you can do to help manage your symptoms.<br><br>Firstly, it is essential that you continue to monitor your symptoms closely, as they can change rapidly. Be sure to take note of any new symptoms that arise or if your current symptoms worsen. Secondly, stay hydrated by drinking plenty of fluids like water, herbal tea, and soup. This will help to keep you hydrated and aid in your recovery.<br><br>I also recommend that you take over-the-counter medication like acetaminophen or ibuprofen to help manage your headache and cough symptoms. Please be sure to follow the recommended dosage and guidelines on the packaging.<br><br>As you are up-to-date with your COVID-19 vaccination, you have some level of protection against the virus. However, it is still essential to isolate yourself from others, wear a mask, and follow proper hand hygiene to prevent the spread of the virus to those around you.<br><br>If your symptoms worsen, or if you experience any difficulty breathing, chest pain, or confusion, please call me immediately. I may need to assess your condition further and determine if additional medical care is necessary.<br><br>I hope these recommendations will help you manage your symptoms and support your recovery. Please do not hesitate to contact me if you have any further questions or concerns. Remember that I am here to support you through this difficult time.<br><br>Take care and stay safe.<br><br>Best regards, |

| | | |
|---|---|---|
| | | [Your Name] |
| ChatGPT4 | Question 9: Hello, I wanted to let you know I tested positive for covid today. I'm having a cough, dry throat, feeling tired, and a small headache. No fevers or aches. I'm up-to-date with my covid vaccine so I'm hoping things won't get worse. I'm trying to rest, doing some nasal rinses, and steam. Anything else specific you'd recommend me to do to treat? Thanks. [PATIENT] | Dear [PATIENT],<br><br>I'm sorry to hear that you're feeling unwell and have tested positive for COVID-19. Your proactive steps in getting vaccinated and reaching out to share your symptoms are commendable.<br><br>Given your current symptoms , cough, dry throat, fatigue, and a small headache , it seems like you have a mild case. As you've already noted, rest is very important at this stage. Make sure you're getting plenty of sleep and are staying well-hydrated. Warm liquids can help soothe the throat and might relieve your cough, so consider having warm water, herbal teas, or broths.<br><br>Over-the-counter medications can be used to manage your symptoms. A simple pain reliever, like acetaminophen (Tylenol), can help with the headache. Be sure to follow the package instructions for any medication you take.<br><br>Monitor your symptoms closely, particularly any changes in breathing. If you notice any worsening, such as difficulty breathing, persistent pain or pressure in your chest, new confusion, inability to wake or stay awake, or bluish lips or face, seek immediate medical attention.<br><br>Regarding your diet, try to maintain a balanced and nutritious one as much as possible. Consuming foods rich in vitamins and minerals will support your immune system. If your appetite is affected, try to eat smaller, more frequent meals instead of three large ones.<br><br>Isolation is crucial to prevent the spread of the virus. The current guidelines recommend staying isolated for at least 5 days from the date of your positive test if you're asymptomatic, or from the onset of symptoms if you're symptomatic, and at least 24 hours have passed without a fever (without the use of fever-reducing medications) and your other symptoms are improving. However, these guidelines may vary by region and the latest local health department guidelines should be followed.<br><br>I'm here to support you throughout your recovery. If you have further questions or if your condition changes, please don't hesitate to reach out. Take care of yourself and rest well.<br><br>Best,<br>[Your Name] |
| CLAIR-Short | Question 9: Hello, I wanted to let you know I tested positive for covid today. I'm having a cough, dry throat, feeling tired, and a small headache. No fevers or aches. I'm up-to-date with my covid vaccine so I'm hoping things won't get worse. I'm trying to rest, doing some nasal rinses, and steam. Anything else specific you'd recommend me to do to treat? Thanks. [PATIENT] | I am sorry to hear you are not feeling well. You need to quarantine for 5 days. If on day 5 you are fever free and your symptoms are improving, then you can come out of quarantine but continue to wear a mask around others for an additional 5 days. You can use over the counter medication to help with your symptoms. Tylenol for fever. Mucinex for cough and congestion. Rest and stay hydrated. If you have any questions or concerns, please let us know. [HCW] |

| | | |
|---|---|---|
| CLAIR-Short | Question 9: Hello, I wanted to let you know I tested positive for covid today. I'm having a cough, dry throat, feeling tired, and a small headache. No fevers or aches. I'm up-to-date with my covid vaccine so I'm hoping things won't get worse. I'm trying to rest, doing some nasal rinses, and steam. Anything else specific you'd recommend me to do to treat? Thanks. [PATIENT] | I am sorry to hear this. Unfortunately, there is not a lot of treatment for covid. The best thing you can do is treat your symptoms. For the cough, you can take plain mucinex. For the dry throat, you can try throat lozenges or warm salt water gargles. For the fatigue, you can try to get as much rest as possible. For the headache, you can take tylenol or ibuprofen. If you have any other symptoms, please let us know. If you don't get better or feel worse, please let us know. Please let us know if there is anything else that we can help you with. Thank you, [HCW] |
| CLAIR-Short | Question 9: Hello, I wanted to let you know I tested positive for covid today. I'm having a cough, dry throat, feeling tired, and a small headache. No fevers or aches. I'm up-to-date with my covid vaccine so I'm hoping things won't get worse. I'm trying to rest, doing some nasal rinses, and steam. Anything else specific you'd recommend me to do to treat? Thanks. [PATIENT] | I am sorry to hear you have COVID. Make sure you drink plenty of fluids. You can also take over the counter cold and flu medicine and tylenol as needed, ( please be mindful of your total acetaminophen intake ; it should not exceed 3, 000 mg per day. ) Avoid NSAIDS ( ibuprofen, naproxen, etc. ) unless you were tak ing these previously for other diagnosis or condition. Continue taking all your prescribed medications, and self - isolate from others for at least 10 days from the beginning of your diagnosis AND until you are fever free for three days without any Tylenol. You can end isolation after this if you are fever - free for 24 hours without the use of fever - reducing medication and your other symptoms have improved. Nevertheless, it is recommended that you continue to wear a mask around others for an additional 5 days. Keep in mind that friend family and household members are considered to be at high risk and should also stay away from you. Keep in mind that friends, family, and household members are considered to be at high risk and should stay away from you. Friends, family, and household members who are healthy adults and at low risk of complications can come into the house wearing a mask for 5 minutes to check on you, help with your care, or bring you food or medicine. Anyone who has been in close contact with you when you were infectious should consider getting tested for COVID - 19. In some cases, your doctor may decide that you need to take an antiviral medication. These medications may help prevent serious problems from occurring. If you start to feel worse, go to the emergency room. When to go to the ER : - You have shortness of breath or difficulty breathing - You have chest pain - You have a fever that does not come down with Tylenol or ibuprofen - You are so drowsy that you are difficult to arouse - New weakness - New numbness - Confusion - New vision changes - New severe headache |
| CLAIR-Long | Question 9: Hello, I wanted to let you know I tested positive for covid today. I'm having a cough, dry throat, feeling tired, and a small headache. No fevers or aches. I'm up-to-date with my covid vaccine so I'm hoping things won't get worse. I'm trying to rest, doing some nasal rinses, and steam. Anything else specific you'd recommend me to do to treat? Thanks. [PATIENT] | Dear Patient,<br><br>Thank you for reaching out to me regarding your recent diagnosis of COVID-19. I understand how frustrating it can be to experience symptoms of illness such as those you described in your letter. I hope you feel better soon. I am sorry to hear about your recent COVID-19 diagnosis. There are several ways to help manage your symptoms at home. Staying hydrated by drinking plenty of water and other fluids is important to prevent dehydration. If you are experiencing a sore throat, gargling with saltwater may help ease discomfort. Over-the-counter medications such as acetaminophen (Tylenol) and ibuprofen (Advil, Motrin) can help reduce body aches and fever. Decongestants can also be taken if you are experiencing congestion, runny nose, or sinus pressure. In general, it is advised to avoid taking aspirin due to its association with increased risk of bleeding complications. It is recommended to continue to monitor your symptoms for any worsening of your condition, including difficulty breathing, chest pain, shortness of breath, loss of consciousness, or confusion. If you experience any of these symptoms, please seek immediate medical attention. Additionally, it is extremely important to remain isolated from others as much as |

| | | |
|---|---|---|
| | | possible until you are no longer experiencing symptoms, or until 5 days after your initial symptom onset, whichever comes first. It is also very important to wear a mask when around others during this time period to help prevent transmission of the virus. If you have questions or concerns, please don't hesitate to reach out to me directly. I will be happy to help in any way I can. Thank you,<br><br>[HCW] |
| Actual Provider Response | Question 9: Hello, I wanted to let you know I tested positive for covid today. I'm having a cough, dry throat, feeling tired, and a small headache. No fevers or aches. I'm up-to-date with my covid vaccine so I'm hoping things won't get worse. I'm trying to rest, doing some nasal rinses, and steam. Anything else specific you'd recommend me to do to treat? Thanks. [PATIENT] | Hi [PATIENT], When did your symptoms start? That will determine if you are eligible for the antiviral, Paxlovid. Best, [HCW] |
